# CRISPR-based environmental detection of *Burkholderia pseudomallei* identifies sanitation gaps and melioidosis risk in northeast Thailand

**DOI:** 10.1101/2024.11.21.24317607

**Authors:** Sukritpong Pakdeerat, Chalita Chomkatekaew, Phumrapee Boonklang, Arin Wongprommoon, Kesorn Angchagun, Yaowaret Dokket, Areeya Faosap, Gumphol Wongsuwan, Premjit Amornchai, Vanaporn Wuthiekanun, Jiramate Changklom, Suwatthiya Siriboon, Parinya Chamnan, Sharon J Peacock, Julian Parkhill, Jukka Corander, Nicholas PJ Day, Nicholas R Thomson, Chayasith Uttamapinant, Somsakul Pop Wongpalee, Claire Chewapreecha

**Author notes:** Correspondence to: Claire Chewapreecha. Contributed equally (Sukritpong Pakdeerat, Chalita Chomkatekaew, Phumrapee Boonklang).

## Abstract

Environmental exposure to *Burkholderia pseudomallei*, the causative agent of melioidosis, remains poorly characterised due to the limited sensitivity of conventional detection methods. This hinders accurate risk mapping and delays public health responses. Here, we developed CRISPR-BEEPs – a sensitive, equipment-light CRISPR-based assay – that demonstrated substantially improved sensitivity (93.5% vs 19.4%) and specificity (100% vs 98.0%) compared to conventional culture-based plate inspection techniques. We applied CRISPR-BEEPs to water samples collected from both natural and piped sources across 15,118 km² in northeast Thailand, including households of confirmed melioidosis patients and controls. *B. pseudomallei* was detected in 73.3% of groundwater samples, 32.4% of surface water, and 28.3% of piped water, with peak detection during the flood season. Importantly, the assay’s improved sensitivity enabled detection of a significant association between environmental *B. pseudomallei* detection within a 10 km radius of participants’ households and melioidosis risk (OR 2.74; 95% CI: 1.38– 5.48) – an association undetectable by conventional methods. These findings highlight major gaps in water treatment and distribution infrastructure and demonstrate the value of high-resolution environmental diagnostics. Strengthening water sanitation and surveillance systems is essential for mitigating melioidosis transmission and addressing the broader burden of waterborne diseases in vulnerable settings.

## Introduction

Access to clean water and adequate sanitation is fundamental to public health^1,2^. The United Nations has issued Sustainable Development Goal 6 (SDG 6) to call for universal access to safe water, improved sanitation, and sustainable resource management. However, in many low-resource settings, water safety remains compromised, increasing the risk of exposure to environmental pathogens that contribute to disease burdens^2–4^. *Burkholderia pseudomallei,* the bacterium that causes melioidosis, can be found in soil and water in tropical and subtropical regions, including South- and Southeast Asia, parts of Africa and the Americas, and northern Australia^5^. Human infection occurs through skin abrasions, inhalation, or ingestion and disproportionately affects individuals with underlying conditions such as diabetes or immunosuppression^6,7^. Melioidosis is a severe infection with a reported case fatality rate of up to 40%^8,9^. Despite its severity, melioidosis remains underdiagnosed and underreported, with environmental pathways through which *B. pseudomallei* spreads are still not fully understood.

Multiple outbreaks have linked *B. pseudomallei* to contaminated water supplies^10–16^. In 2023, a cluster of ten neurological melioidosis cases in southern India – eight of which were fatal – was traced to contaminated saline used in a dental clinic^17^. In 2021, an outbreak at a COVID-19 field hospital in Thailand resulted in 25 confirmed cases and eight deaths, primarily among patients treated with corticosteroids^11^. The source was traced to inadequately chlorinated water used for daily hygiene. Similarly, outbreaks in northern^13,16^ and western^12^ Australia have been associated with unchlorinated or poorly maintained water systems, collectively resulting in 15 confirmed cases and nine deaths. A notable outbreak in the United States, involving four confirmed cases and two deaths, was traced to a contaminated aromatherapy spray imported from India^14^, highlighting a global relevance of water sanitation in preventing melioidosis – even outside endemic regions. Although chlorination is generally effective against *B. pseudomallei*, the bacterium can survive within biofilms and may display moderate chlorine resistance^18,19^. The World Health Organisation (WHO) water safety framework emphasises the need for multilayered strategies – such as maintaining effective disinfection, preventing environmental contamination, and mitigating biofilm development within distribution systems – to minimise the risk of *B. pseudomallei* contamination^20^. However, implementation of these measures remains challenging in many endemic regions^3,21^. Moreover, current water monitoring practices, which typically rely on faecal indicator bacteria such as *Escherichia coli*^22^, are insufficient for identifying non-fecal pathogens like *B. pseudomallei*, which can persist and replicate in water systems independently of faecal contamination.

Environmental surveillance of *B. pseudomallei* has traditionally depended on culture-based methods using selective media followed by visual inspection before confirmation of suspected colonies with biochemical or serological assays^23–25^. While these methods have provided a foundation for detection, they suffer from low sensitivity and are increasingly challenged by the presence of antibiotic-resistant environmental organisms that complicate visual inspection. Molecular techniques such as PCR offer improved sensitivity and specificity, but their use in field settings is limited. PCR reactions are sensitive to environmental inhibitors^26,27^, require extensive protocol optimisation, and depend on specialised equipment and trained personnel. To overcome these limitations, emerging CRISPR-based diagnostics technologies – such as Cas12 and Cas13-based detection systems – show considerable promise^28,29^. When combined with isothermal amplification methods like recombinase polymerase amplification (RPA) or loop-mediated isothermal amplification (LAMP), they form powerful platforms such as SHERLOCK (Cas13 with RPA)^30–32^ and DETECTR (Cas12 with either RPA^33,34^ or LAMP^35^). These integrated systems have demonstrated high sensitivity and specificity in clinical diagnostics and are increasingly being explored for environmental surveillance^34^. Their key advantages include tolerance to common environmental inhibitors, minimal instrumentation requirements, and suitability for deployment in low-resource or field settings^36,37^.

In this study, we developed and evaluated a sensitive, equipment-light, CRISPR-based assay – originally designed for clinical diagnostics^38,39^ – for its potential use in environmental surveillance of *B. pseudomallei*. The assay is based on DETECTR^33^ technology, combining RPA with CRISPR-Cas12-mediated detection. Upon recognising the target sequence, the Cas12 enzyme activates collateral cleavage of a reporter molecule, generating a detectable signal that can be read using a simple lateral flow dipstick – making it suitable for low-resource settings. We further optimised this assay for environmental applications by developing a new laboratory protocol, which we termed **CRISPR-BEEPs** (**CRISPR**-based ***B****. pseudomallei* **E**nhanced **E**nvironmental **P**late **s**weep sampling). This method was implemented across 15,118 km^2^ region in northeast Thailand, an area endemic for melioidosis. Our study included diverse water source, such as piped water, groundwater, and natural surface waters. In addition to evaluating analytical and field performance, we assessed the association between environmental *B. pseudomallei* detection and melioidosis incidence. CRISPR-BEEPs provided markedly improved sensitivity over conventional culture, enabling more accurate exposure risk assessment and reinforcing the need for enhanced water sanitation and access to clean water in endemic regions.

## Results

### Environmental microbial diversity in the tropics and subtropics

To apply DNA-based detection methods - originally designed for clinical applications - for environmental surveillance, we first evaluated the *in silico* specificity and sensitivity of selected *B. pseudomallei* DNA targets (**Figure 1a**) within the context of highly diverse microbial communities found in tropical and subtropical environments. Water sources in melioidosis-endemic areas (**Supplementary Figure 1**) are ecologically complex^40^, containing fast-growing fungi, diverse bacterial taxa (including both fast- and slow-growing species), and various other eukaryotic microbes. These diverse microbial communities present analytical challenges for the specific and sensitive detection of *B. pseudomallei*. To characterise the microbial environment where *B. pseudomallei* resides, we analysed metagenome-assembled genomes (MAGs) from the Earth Microbiome Project^41^ – a global dataset representing microbial communities from freshwater, wastewater, soil, plant rhizospheres, fungi, and human and animal faeces. Focusing specifically on MAGs originating from tropical (23.5°N - 23.5°S) and subtropical (23.5° – 40° in both hemispheres) regions that overlap with the established melioidosis-endemic areas^5^ (**Figure 1b**), we compiled a dataset comprising 27,771 MAGs representing 9,739 operational taxonomic units (OTUs). This dataset reflects the broad microbial diversity potentially co-existing with *B. pseudomallei* in these environments.

**Figure 1.**
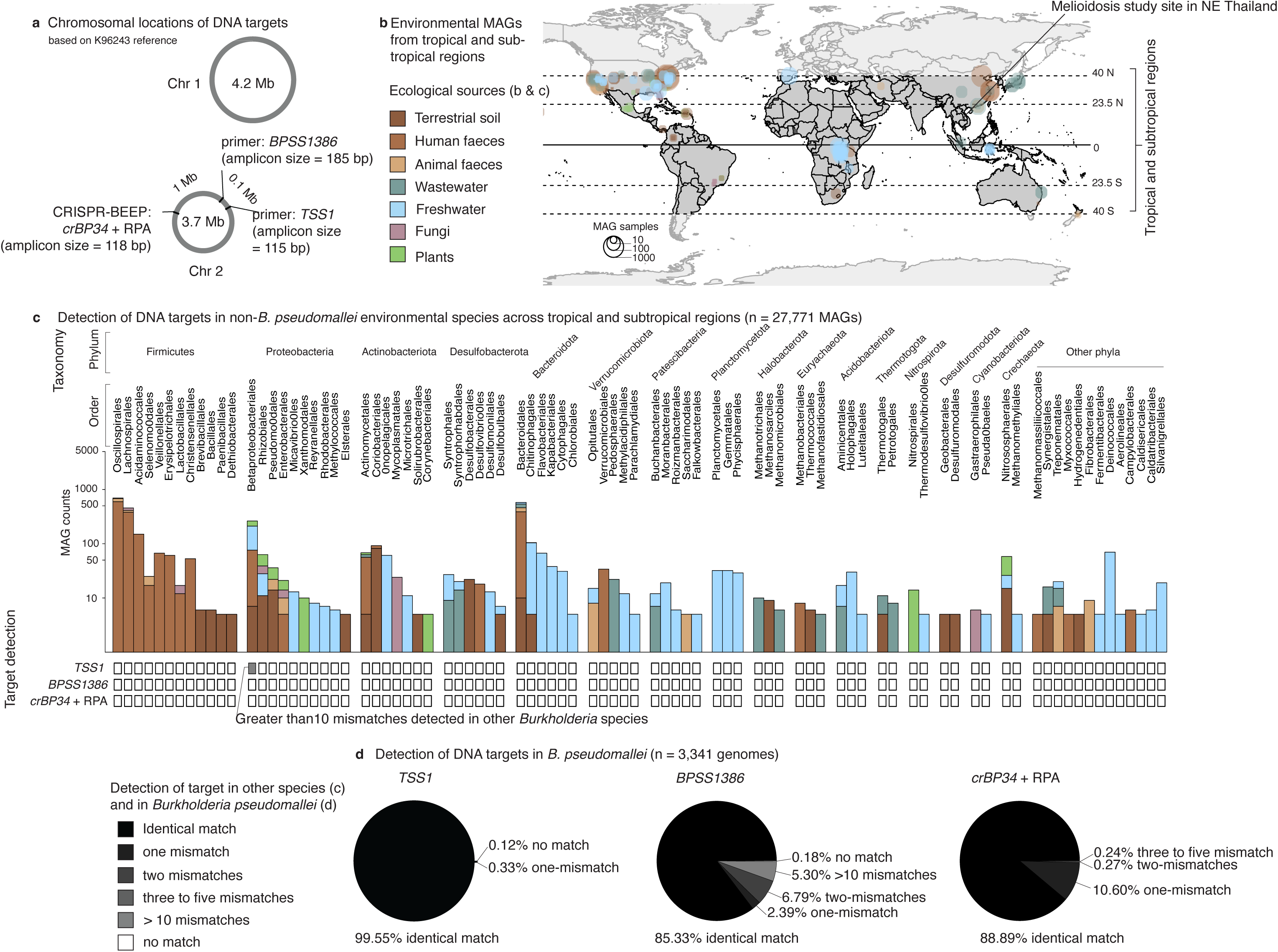
Molecular targets for *B. pseudomallei* detection and their *in silico* specificity and sensitivity in the context of environmental microbial background. (a) DNA targets include two qPCR primer sets (*TTS1* and *BPSS1386*) and the CRISPR-BEEPs locus (RPA primers and *crBP34*), located in distinct genomic regions for independent detection. (b) Data availability of 27,771 metagenome-assembled genomes (MAGs) from tropical and subtropical regions, representing microbial communities from soil, freshwater, wastewater, feces, fungi, and plants. (c) Taxonomic abundance of environmental MAGs found in (b) and assessment of partial sequence matches between these taxa and the DNA targets shown in (a). (d) Conservation of DNA targets in the global *B. pseudomallei* population, based on 3,341 high-quality genome assemblies. In (b) and (c), MAGs are colour-coded by source. In (c) and (d), matching status is visualized using a greyscale gradient: black indicates perfect matches, grey indicates partial mismatches, and white indicates no match.

Among freshwater samples, 50.3% of MAGs were dominated by the phyla Proteobacteria (473/2644, 17.9%), Bacteroidota (471/2644, 17.8%), and Patescibacteria (333/2644, 12.6%). In contrast, microbial communities from faecal-contaminated and soil-associated sources were primarily composed of the phyla Firmicutes (2140/4467, 47.9%), Bacteroidota (775/4467, 17.3%), and Proteobacteria (417/4467, 9.3%) (**Figure 1c**). The consistent presence of Proteobacteria and Bacteroidota across water, soil, faeces, and rhizosphere environments highlights their ecological ubiquity. Notably, Proteobacteria includes the genus *Burkholderia*, which comprises a range of ecologically and clinically important species. These include non-pathogenic environmental bacteria such as *Burkholderia thailandensis*; plant pathogens such as *Burkholderia glumae*, and *Burkholderia plantarii*; and opportunistic human pathogens including *Burkholderia cepacia* complex and *Burkholderia pseudomallei*. However, *B. pseudomallei* itself was not detected in any of the MAG datasets analysed. This is likely due to a combination of factors: (1) the bacterium’s typically low abundance in environmental samples, (2) insufficient sequencing depth and biases in MAG assembly or taxonomic classification, and (3) the masking of rare *B. pseudomallei* sequences by the high background of other microbial and archaeal DNA.

### *In silico* specificity and sensitivity of *B. pseudomallei* DNA targets

The challenges outlined above mirror real-world difficulties in detecting *B. pseudomallei* DNA within complex environmental samples containing high-background levels of non-target microbial, archaeal, and closely related *Burkholderia* species DNA. To evaluate assay performance in such conditions, we assessed the *in silico* specificity and sensitivity of three molecular targets: *TTS1*^42^ and *BPSS1386*^43^ (used in qPCR reference test), and the CRISPR-BEEPs target region, which includes the guide RNA *crBP34* and its associated RPA primers^38^ (**Figure 1a** & **Table S1**).

When mapped against the 27,771 environmental MAGs, the CRISPR-BEEPs target region (*crBP34* + RPA primers) and *BPSS1386* qPCR primers showed no sequence matches, indicating high specificity (**Figure 1c**). In contrast, *TTS1* primers exhibited partial matches to other *Burkholderia* species, with up to 10 mismatches – most critically at the 3’ end of the primer, where mismatches are known to compromise PCR efficiency^44,45^. Nonetheless, all three targets are sufficiently specific to *B. pseudomallei* in the context of available MAG data, with a low likelihood of off-target amplification in environmental samples.

To assess sequence conservation within the *B. pseudomallei* population, we screened 3,341 high-quality *B. pseudomallei* genome assemblies^46–50^ representing the species’ global diversity (**Figure 1d**). Exact matches to the target regions were found in 99.55% of genomes for *TTS1*, 85.33% for *BPSS1836*, and 88.89% for the CRISPR-BEEPs crBP34 target and its associated RPA primers. Given that PCR^44,45^, RPA^51^, and CRISPR-cas12a^52^ assays can tolerate a small number of mismatches – particularly outside critical regions such as the 3’ end of primers or the seed region of crRNAs – we applied a conservative threshold allowing up to two nucleotide differences. Under this relaxed criterion, the predicted coverage increased to 99.88% for *TTS1*, 94.52% for *BPSS1386*, and 99.76% for CRISPR-BEEPs’ target regions. These results confirm that the three targets are both conserved in *B. pseudomallei* and are unlikely to cross-react with non-target organisms, thereby validating their use in molecular assays for environmental surveillance.

### Analytical sensitivity of molecular assays and comparison to environmental *B. pseudomallei* levels

To determine the analytical sensitivity of the CRISPR-BEEPs assay and compare it with standard qPCR methods, we created a simulated environmental DNA background. Genomic DNA from *B. pseudomallei* was spiked into a constant background of DNA extracted from bacteria commonly found in tropical freshwater and piped water systems^41,53^, including *Escherichia coli*, *Klebsiella pneumoniae*, *Enterobacter cloacae*, *Serratia marcescens*, *Citrobacter freundii*, and *Leclercia adecarboxylata*. The concentration of *B. pseudomallei* DNA was serially diluted while background DNA was held constant (**Figure 2a-c**). Using this matrix, qPCR detected *B. pseudomallei* at 5 genomic copies/μL with *TTS1* primers and 10 copies/μL with *BPSS1386* primers. The CRISPR-BEEPs assay, using a lateral flow dipstick readout, achieved a detection limit of 20 copies/μL. While all three assays demonstrated high sensitivity, these detection thresholds remain higher than the *B. pseudomallei* concentrations typically found in environmental water samples^54–57^.

**Figure 2.**
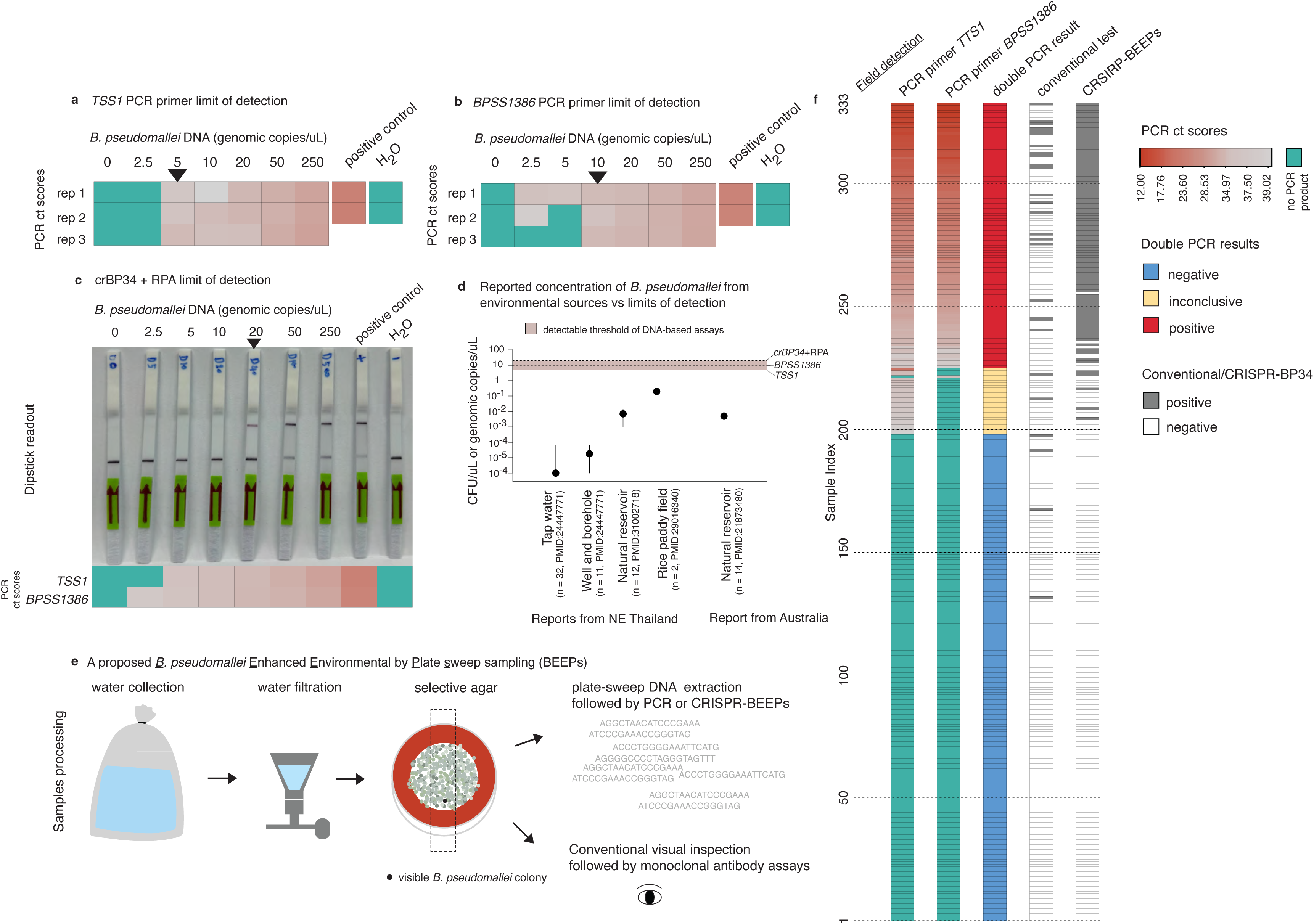
Analytical and field performance of CRISPR-BEEPs and qPCR, and proposed laboratory protocol to improve detection. (a - c) Analytical sensitivity of qPCR assays using *TSS1* (a), *BPSS1386* (b), and crBP34 + RPA with lateral flow readout (c). Ct values are colour-coded: green = no amplification, red gradient = increasing signal intensity (dark red = low Ct, pale red = high Ct). Black triangles indicate the lowest concentration at which three replicates were consistently detected. (d) Comparison of assay detection limits (in genomic copies/µL) with reported concentrations of *B. pseudomallei* in environmental water samples (in CFU/µL), assuming a 1:1 genome-to-CFU ratio. (e) Proposed laboratory workflow for environmental surveillance, incorporating culture enrichment to ensure sufficient bacterial load for detection and a plate-sweep approach to reduce subjectivity from colony selection. (f) Field sensitivity and specificity of the protocol in (e), comparing qPCR classification (using both *TSS1* and *BPSS1386*), conventional culture-based detection, and CRISPR-BEEPs (n samples = 333).

Field studies in melioidosis-endemic regions have consistently shown that *B. pseudomallei* is present in water sources at low levels: a median of 1 x 10^-6^ CFU/µL (range: < 0.1 – 6.3 10^-5^ CFU/µL) in public piped water^54^; 1.8 x 10^-5^ CFU/µL (range: < 0.1 – 6.5 x 10^-5^ CFU/µL) in household wells and borehole^54^; 7.0 x 10^-3^ CFU/µL (range: 0.1 – 1.4 x 10^-2^ CFU/µL) in natural reservoirs in northeast Thailand^55^; and 5.0 x 10^-3^ CFU/µL (range: 0.01 – 1.13 x 10^-1^ CFU/µL) in Australia water sources^56^, with the highest concentration reported at 0.25 CFU/ µL in rice paddies^57^. Although *B. pseudomallei* may exist in a viable but non-culturable state, and not all DNA detected reflects viable organisms, we conservatively assume a 1:1 equivalence between colony-forming units (CFU) and genomic copies to benchmark assay performance (**Figure 2d**). On this basis, most environmental water samples are likely to contain *B. pseudomallei* at concentrations below the direct detection limits of current molecular assays, underscoring the necessity of pre-enrichment to improve detection sensitivity. To overcome this limitation, we incorporated a selective enrichment step using Ashdown’s medium (**Figure 2e**). This approach enhances the proliferation of *B. pseudomallei* while suppressing competing microorganisms, thereby increasing genomic copy number to the levels within the detectable range of both qPCR and CRISPR-BEEPs assays.

### Field sensitivity and specificity of CRISPR-BEEPs for *B. pseudomallei* detection in environmental water samples from northeast Thailand

To evaluate the real-world performance of CRISPR-BEEPs in environmental surveillance, we applied the assay to water samples collected across northeast Thailand, a region hyperendemic for melioidosis. Between November 2020 and November 2021, 356 water samples were obtained from various sources, including household piped water, private boreholes, and natural water reservoirs^49^ (**Figure 3a & c**, **Supplementary Figure 1**). Following filtration, all samples were subjected to selective culture using Ashdown’s agar (**Figure 2e**). Due to fungal overgrowth, 21 samples were excluded, resulting in 335 interpretable cultures. Initial identification was performed using conventional culture-based methods and visual inspection of colony morphology, with suspected colonies further tested using monoclonal antibodies^58,59^. Out of 87 visually suspicious colonies, only 23 were confirmed as *B. pseudomallei* (**Supplementary Figure 2a & b**). This highlights the known limitations of culture-based detection. *B. pseudomallei* often grows slowly and can be overgrown by faster-growing environmental microbes, while colony morphology alone lacks sufficient specificity for accurate identification.

**Figure 3.**
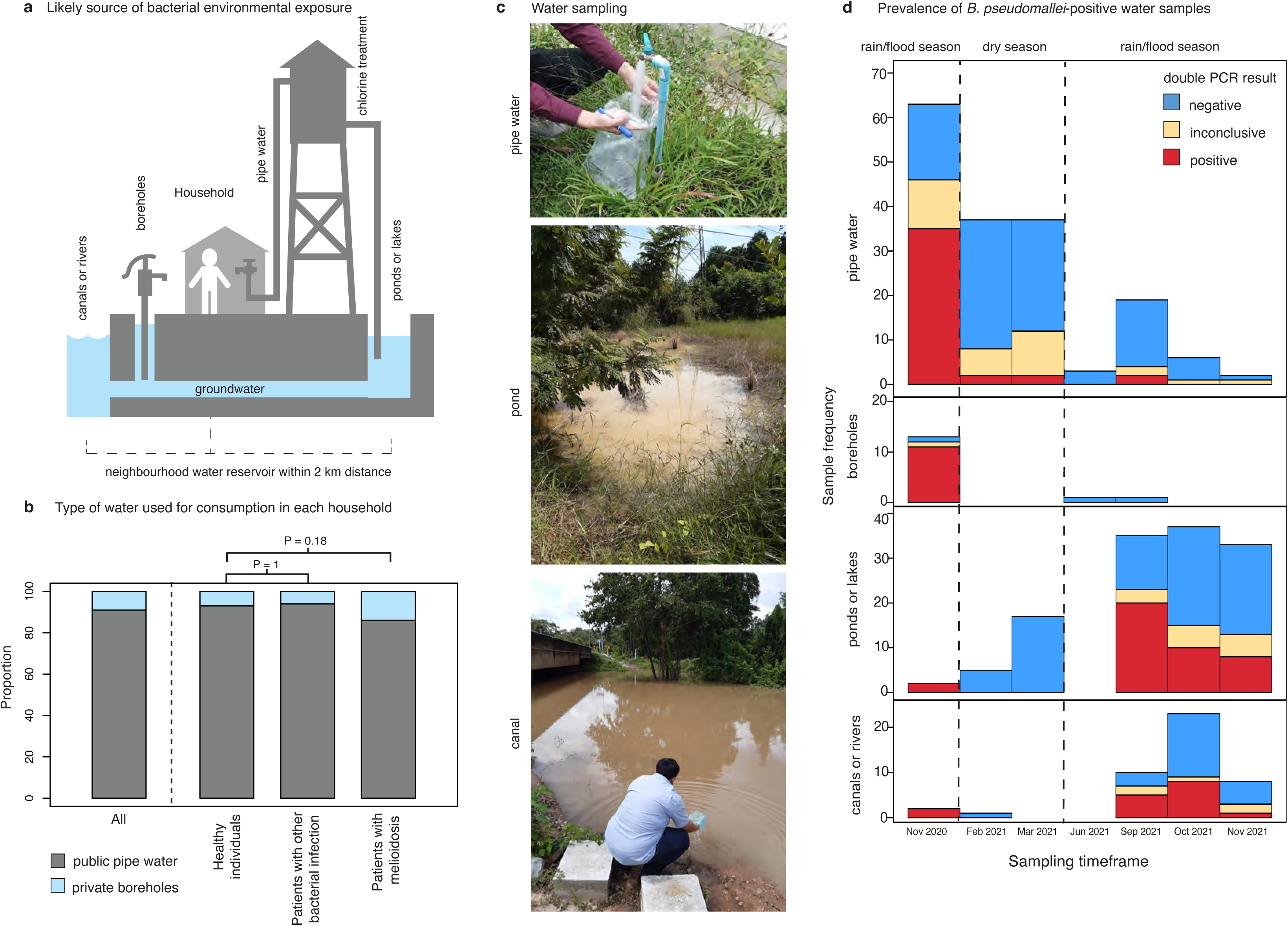
Water sources and seasonal variation in *B. pseudomallei* detection. (a) Diagram of water sources: groundwater (boreholes), surface water (ponds, lakes, canals, rivers) and piped water system. (b) Proportion of participants (n = 1,132) reporting use of public piped water and boreholes, grouped by clinical outcome. (c) Representative sampling locations: household piped water (top), stagnant water from a pond (middle), and flowing water from a canal (bottom). Gloves were not worn inside household due to cultural practices; however, contamination from human microbiota is unlikely due to selective enrichment of *B. pseudomallei.* (d) Seasonal variation in detection outcomes (positive = red, inconclusive = yellow, negative = blue) across water sources: piped water (n = 145), boreholes (n = 15), ponds or lakes (n = 129), and canals or rivers (n = 44).

To overcome these constraints, we implemented molecular detection using a plate-sweep approach: all colony growth from Ashdown agar plates was harvested en masse for DNA extraction, irrespective of colony appearance (**Supplementary Figure 2c**). High-quality microbial DNA was successfully extracted from 333 of the 335 enriched cultures. Each DNA sample was screened using three molecular targets – *TTS1* and *BPSS1386* primers (qPCR), and *crBP34* with its RPA primers (CRISPR-BEEPs) – as described previously. Given its higher analytical sensitivity (**Figure 2a-c**), qPCR was used as the reference standard. Samples were classified as *B. pseudomallei*-positive if both qPCR targets were detected (“double-qPCR positive”, n = 108), inconclusive if only one target was detected (n = 27), and negative if neither was detected (n = 198) (**Figure 2f**).

CRISPR-BEEPs showed high concordance with this classification, correctly identifying 101 of 108 double-qPCR positive samples (sensitivity 93.52%, 95% CI: 87.10 – 97.35%) (**Table 1**). In contrast, the conventional culture-based method detected only 21 of these 108 samples (sensitivity 19.44%, 95% CI: 12.46 – 28.17%). CRISPR-BEEPs also demonstrated 100% specificity, correctly identifying all qPCR-negative samples (100.00%, 95% CI: 98.15 – 100.00%), while the conventional method showed slightly lower specificity at 97.98% (95% CI: 94.91 – 99.45%). Statistical comparison using McNemar’s test (p-value = 5.83 x 10^-8^) confirmed the significantly improved diagnostic performance of CRISPR-BEEPs compared to conventional plate inspection approach. These results demonstrate that CRISPR-BEEPs, which integrates selective enrichment and plate-sweep DNA extraction, offers a highly sensitive, specific, and scalable approach for detecting *B. pseudomallei* in complex environmental samples, enabling more accurate and reliable risk assessments in melioidosis-endemic regions.

**Table 1.** Improved sensitivity offered by CRISPR-BEEPs compared to conventional approach.

|  | Sensitivity<br>(percent, 95% confidence interval) | Specificity<br>(percent, 95% confidence interval) |
| --- | --- | --- |
| Conventional plate inspection | 21 of 108<br>(19.44%, 12.46-28.17) | 194 of 198<br>97.98% (94.91-99.45) |
| CRISPR-BEEPs | 101 of 108<br>(93.52%, 87.10-97.35) | 198 of 198<br>(100%, 98.15-100.00) |

### Enhanced environmental resolution reveals the true burden of *B. pseudomallei* in northeast Thailand, including piped water system

To better characterise the environmental distribution of *B. pseudomallei*, we analysed the type-specific and seasonal patterns of bacterial detection across a range of water types in northeast Thailand, using double-qPCR positivity as the reference standard (**Figure 3**). *B. pseudomallei* was detected in all major water types sampled, including public piped water (41/145 samples, 28.3%), surface water from both stagnant (ponds and lakes) and flowing (canals and rivers) bodies (56/173 samples, 32.4%), and groundwater from private boreholes (11/15 samples, 73.3%). Among these, groundwater exhibited the highest positivity rate. This elevated positivity in borehole samples is consistent with prior studies^60,61^, supporting the hypothesis that subsurface aquifers – shielded from sunlight, drying, and temperature fluctuation – may act as stable reservoirs for the bacterium. Detection of *B. pseudomallei* in both private boreholes and public piped water is particularly concerning, given that participant interviews confirmed these sources are commonly used for everyday household activities such as bathing, food preparation, and dishwashing (**Figure 3b**). This underscores a direct route of potential exposure in endemic settings.

Seasonality also played a substantial role in *B. pseudomallei* detection rates (**Figure 3d, Supplementary Figure 3**). During the dry season, surface water bodies often receded or dried up completely, limiting both sample availability and pathogen detectability. In contrast, the rainy and flood seasons brought increased precipitation and widespread surface runoff^62^, which may mobilise *B. pseudomallei* from deeper soil layers into water bodies and man-made systems, thereby elevating exposure risk. This pattern was clearly reflected in our findings (**Figure 3d**): 56 of 144 (38.9%) surface water samples collected during the wet season were positive, compared to none of 23 dry-season samples (χ^2^ p = 5.75 x 10^-4^). A similar seasonal trend was observed in public piped water, where positivity increased from 5.3% in the dry season (4/63 samples) to 45.1% during the rainy or flood seasons (37/82 samples, χ^2^p < 2.2 x 10^-16^). These high-resolution, year-round surveillance data confirm and extend earlier multi-year observations^60,61^, showing that wet-season conditions substantially increase the environmental burden of *B. pseudomallei* and likely elevate the risk of human exposure through contaminated water sources.

### Environmental exposure and risk of melioidosis

The relationship between *B. pseudomallei* in environmental sources and the incidence of melioidosis has primarily been described qualitatively, with limited population-level quantitative evidence. This gap stems in large part from the historically low sensitivity of environmental detection methods^63^. Using our improved protocol for detecting *B. pseudomallei* in water sources (**Figure 2f**), we investigated whether individuals who either directly used these contaminated water sources or lived in close proximity to the sampling sites subsequently developed melioidosis. Participants were classified into three outcome groups: confirmed melioidosis cases, patients with other bacterial infections, and healthy controls with no prior history of melioidosis. All groups resided within the same endemic regions, allowing for comparison of environmental exposure under similar ecological and environmental conditions (**Figure 4a**, **Figure 5a** & **b**).

**Figure 4.**
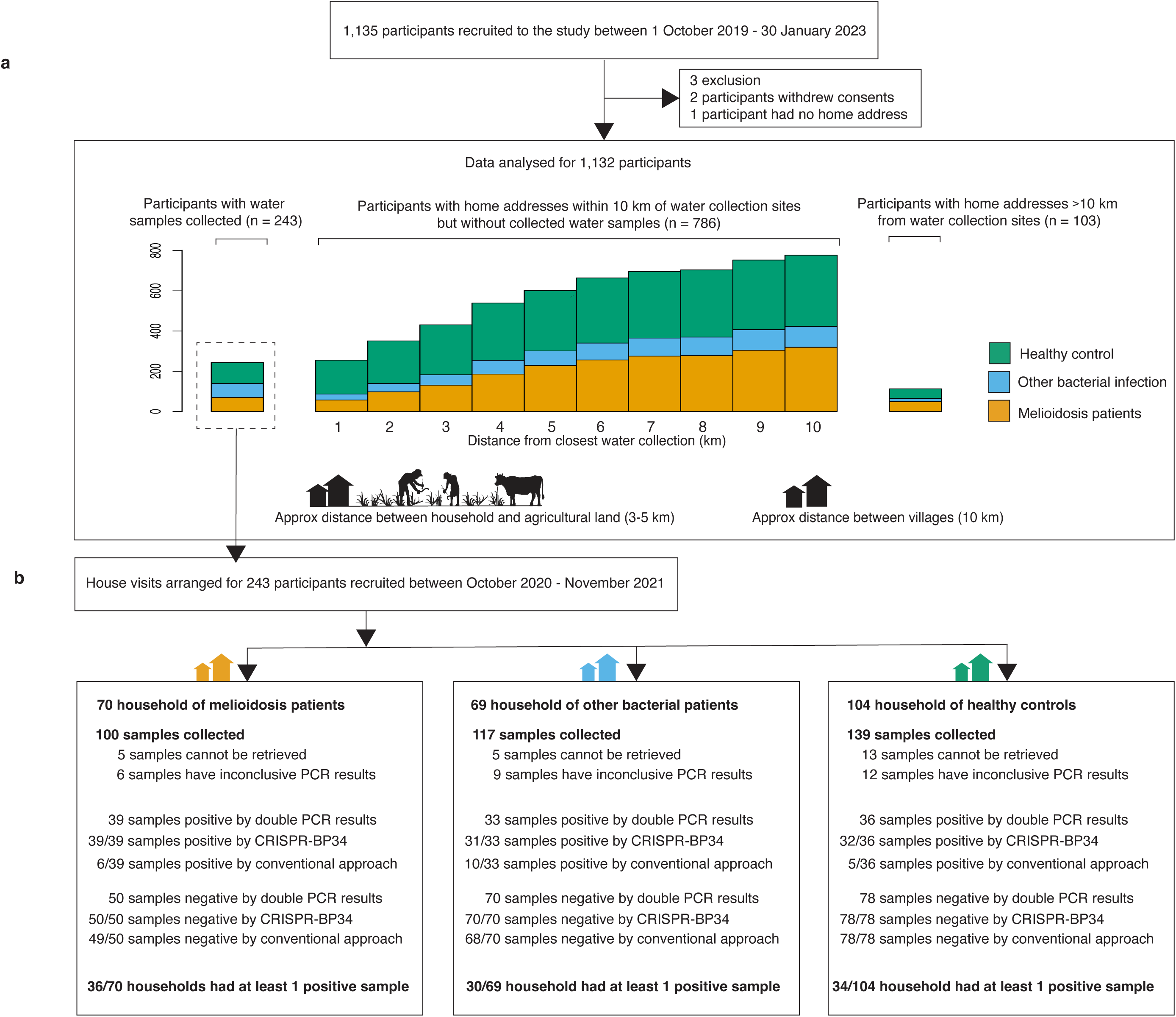
The study flow and data distribution. (a) Cohort overview. Stacked bar plots summarise participants enrolled across time period: those with direct water sampling between November 2020 and November 2021, and additional participants from 2019-2020 and 2021-2023 who lived within 1-10 km of the sampling sites but without direct water collection. Participants are grouped by clinical status: clinically-confirmed melioidosis (orange), other bacterial infections (blue), and healthy controls with no history of melioidosis (green). The 3-5 km radius corresponds to typical travel distance for villager agricultural work, while 10 km approximates the distance between neighbouring villages. (b) Detection of *B. pseudomallei* in household water samples from the cohort by assay type. For household with multiple water samples, a household was considered positive if at least one sample tested positive.

**Figure 5.**
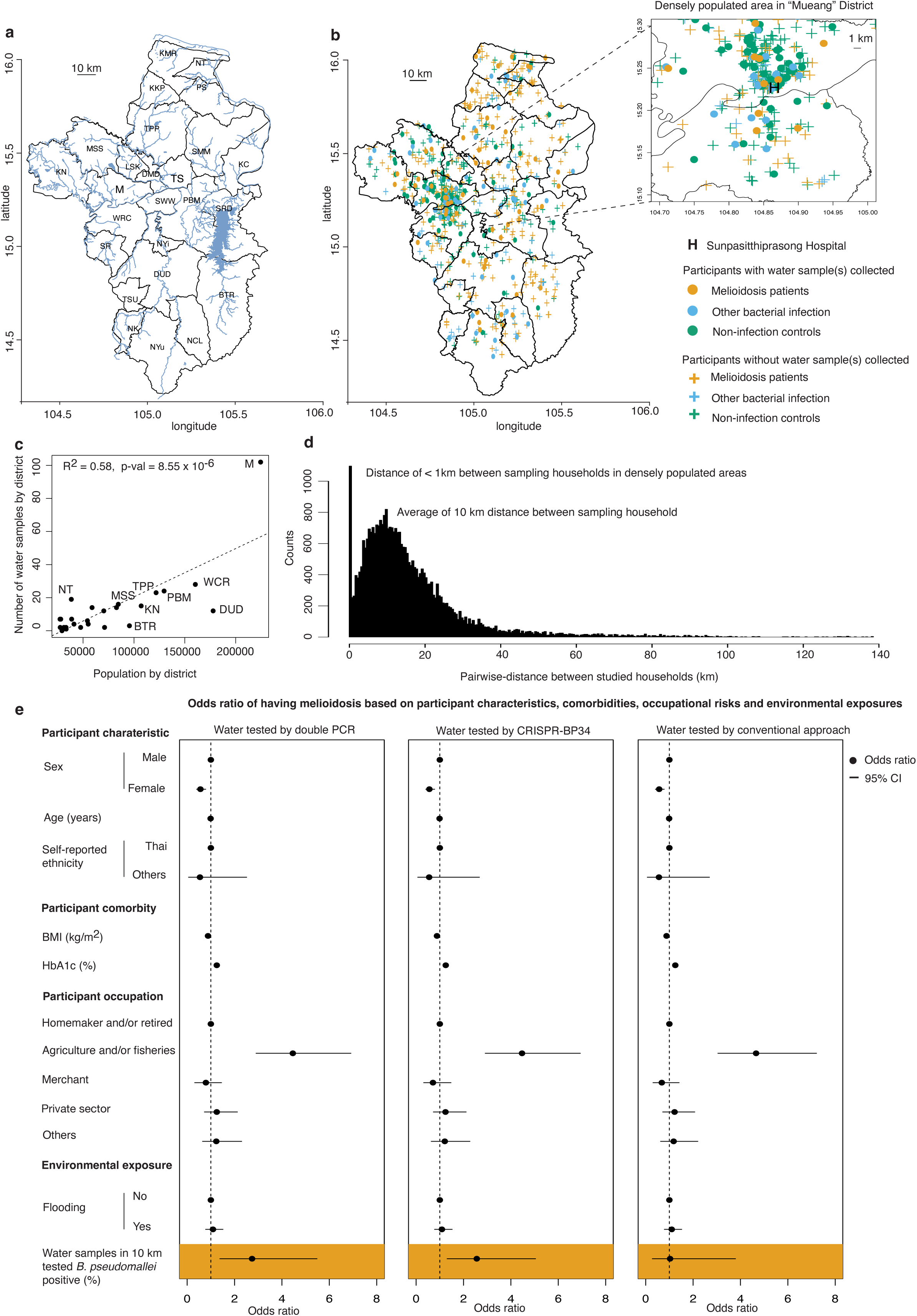
Association between B*. pseudomallei* detection in water sources and melioidosis incidence. (a) Geographical map of Ubon Ratchathani, a province in northeast Thailand, where 1,012 of the 1,135 study participants reside, highlighting local water systems and terrain. (b) Locations of participant households, including those with directly collected water samples and those without samples but residing in close proximity. (c) Positive correlation between the number of water samples collected and population density per district. (d) average distance (in kilometres) between participant households in the cohort. (e) Adjusted odds ratios from multivariable logistic regression showing association between melioidosis and *B. pseudomallei* detection by double-qPCR, CRISPR-BEEPs, and the conventional culture-based plate inspection approach. Districts are abbreviated: Buntharik (BTR), Don Mot Daeng (DMD), Det Udom (DUD), MSS, Khong Chiam (KC), Kut Khaopun (KKP), Khemarat (KMR), Khuang Nai (KN), Lao Suea Kok (LSK), Mueang (M), Muang Sam Sip (MSS), Na Chaluai (NCL), Nam Khun (NK), Na Tan (NT), Na Yia (NYi), Nam Yuen (NYu), Phibun Mangsahan (PBM), Pho Sai (PS), Samrong (SR), Sawang Wirawong (SWW), Sirindhorn (SRD), Si Mueang Mai (SMM), Trakan Phuet Phon (TPP), Tan Sum (TS), Thung Si Udom (TSU), Warin Chamrap (WRC)

A total of 243 participants enrolled between 2020 and 2021 underwent household-level water sampling within three months of either melioidosis diagnosis (for cases) or recruitment (for controls), providing a temporally relevant assessment of exposure (**Figure 4b**, **Supplementary Figure 4**). *B. pseudomallei* was detected in household water sources of 36 out of 70 melioidosis patients (51.4%), significantly higher than the detection rate among healthy controls (34/104 households, 32.7%, Fisher’s exact test p = 0.04). However, this prevalence was not significantly different from that observed in patients with other bacterial infections (30/69 households, 43.5%, p = 0.6). Univariable regression analyses revealed that melioidosis patients had a higher prevalence of established risk factors such as diabetes mellitus and agricultural occupation when compared to patients with other bacterial infections (**Supplementary tables 2-3**). These findings reinforce the notion that while environmental exposure is a necessary component, it is not sufficient to cause disease on its own. Host susceptibility, particularly underlying conditions and occupational exposure, plays a role in determining who progresses from environmental contact to clinical infection.

### Combined effect of environmental and host factors

To investigate the interplay between host factors and environmental exposure, we combined data from two subgroups: 243 participants with direct household-level sampling and 889 participants without direct water sampling but residing in the same endemic region. Together, these groups comprised a cohort of 1,132 participants enrolled between 2019 and 2023 (**Figure 4a**, **Figure 5a & b**). Water samples were systematically collected across all districts, with sampling density scaled to population distribution (**Figure 5c**, R^2^ = 0.58, p = 8.55 x 10^-6^). The average distance between sampling locations was approximately 10 km, corresponding to a typical distance between villages in the study region (**Figure 5d**). In more densely populated areas, such as the central “Mueang” district, sampling was more frequent to ensure adequate environmental representation relative to the population density. Collectively, this provided a spatial proxy for estimating local exposure to *B. pseudomallei*.

Multivariable logistic regression analysis was conducted using all covariates that were significant in univariate models, along with *B. pseudomallei* detection at multiple spatial scales – from household level up to 10 km from a participant’s residence (**Figure 5e**, **Supplementary Tables 4-13**). Several factors emerged as independent predictors of melioidosis. Elevated HbA1c levels, indicating poor glycemic control, were strongly associated with increased disease risk (adjusted OR 1.25, 95% CI 1.19-1.31, p < 0.001). Occupation also played a major role: individuals working in agriculture, fisheries, or farming had significantly higher odds of infection (adjusted OR 4.46, 95% CI 2.91-6.91, p < 0.001), consistent with greater environmental exposure. In contrast, female sex (adjusted OR 0.56, 95% CI 0.40-0.78, p < 0.001), and being overweight or obese (adjusted OR 0.88, 95% CI 0.84-0.91, p < 0.001) were associated with a lower risk of melioidosis. These protective associations likely reflect reduced participation in high-exposure occupation, particularly compared to lean male agricultural workers who are more frequently exposed to environmental sources of *B. pseudomallei*.

After adjusting for host-related risk factors, environmental detection of *B. pseudomallei* emerged as an independent predictor of melioidosis incidence. Using qPCR-based detection methods, a significant association was observed when environmental *B. pseudomallei* was detected within a 7 km radius of participant’s household (adjusted OR 1.91, 95% CI 1.09-3.34, p = 0.024, **Supplementary Table 7**). CRISPR-BEEPs identified a similar association at a 9 km radius (adjusted OR 1.87, 95% CI 1.01-3.5, p = 0.048, **Supplementary Table 5**). The association strengthened further at a 10 km radius – a distance corresponding approximately to the administrative area of a typical rural village (**Figure 5e, Supplementary Table 4**). At this scale, qPCR detection of *B. pseudomallei* was associated with an adjusted OR of 2.74 (95% CI 1.38-5.48, p = 0.004), while CRISPR-BEEPs detection yielded an adjusted OR of 2.55 (95% CI 1.31-5.84, p = 0.006). In contrast, conventional culture-based plate methods failed to detect any significant relationship between environmental positivity and melioidosis incidence. These findings highlight the importance of sensitive molecular diagnostics for identifying high-risk environmental exposure and support village-level risk mapping as a strategy for targeted intervention.

### Discussion and call for improved water sanitation

Our study provides the first statistical evidence linking the presence of *B. pseudomallei* in household and community water sources to clinical cases of melioidosis, a connection previously suspected but not demonstrated with confidence. Crucially, this association became apparent only through the use of sensitive molecular diagnostics. Conventional detection methods – such as culture-based plate inspection commonly used in prior studies^63^ – frequently failed to detect *B. pseudomallei* in environmental samples, likely due to their limited sensitivity and high false-negative rates (**Figure 2f**). In contrast, molecular tools such as double-qPCR and the CRISPR-BEEPs assay revealed strong associations between environmental exposure and melioidosis risk, particularly through household piped water. The CRISPR-BEEPs assay is promising. Its high sensitivity and specificity, combined with minimal equipment requirements, make it suitable for resource-limited settings where melioidosis is endemic. Its deployment at scale could offer early warning of environmental risk and guide timely interventions.

While the study offers new insights, several limitations must be acknowledged. First, overgrowth of fungi during culture enrichment led to the loss of some samples. In rare samples with high *B. pseudomallei* concentrations (>5 genomic copies/µL, **Figure 2d**), direct nucleic acid detection may bypass this issue and enable detection. However, this approach carries the risk of detecting DNA from non-viable organisms, potentially leading to an overestimation of true environmental exposure and an exaggerated association between environmental presence and disease. To balance sensitivity with biological relevance, we retained the culture enrichment step to selectively amplify *B. pseudomallei*, ensuring that the bacterial load reached a detectable threshold and was disease-relevant.

Second, a subset of double-qPCR results was inconclusive. Although the primers were validated both *in silico* in this study and *in vitro* in previous work^42,64^, we cannot fully exclude the possibility of off-target amplification, particularly involving other *Burkholderia* species that are more abundant in the environment. This limitation arises in part from the lack of comprehensive environmental genomic data from the study region, which reduces confidence in excluding potential cross-reactivity with non-target taxa. Additionally, some inconclusive results may reflect limited primer coverage across the genetically diverse *B. pseudomallei* population or suboptimal binding efficiency (**Supplementary Figure 5**). To mitigate these issues and improve analytical robustness, we required concordant results from two independent genomic targets for confirming both positive and negative detections. This dual-target approach minimises false positives, which is particularly important when working with complex environmental samples where horizontal gene transfer is possible and microbial diversity remains under-characterised.

Third, environmental sampling coverage was incomplete: only 243 of 1,132 participants had household water samples collected with a total of 356 samples. This limitation, primarily due to SARS-CoV-2-related travel restrictions and logistical constraints, limits full spatial analysis. Nevertheless, the detection of *B. pseudomallei* in water samples from 2020–2021 suggests the presence of a persistent environmental reservoir, potentially in deep soil or groundwater, that may continue to contaminate surface and piped water sources. While this interpretation should be made with caution, it aligns with the distribution of clinical melioidosis cases observed from 2019 to 2020, and from 2021 to 2023.

Importantly, this study implicates piped water as a potential transmission route—highlighting the public health consequences of inadequate water sanitation. In many areas, insufficient chlorine dosing fails to neutralise pathogens, while ageing infrastructure, pipe breaks, and poor maintenance allow recontamination of treated water. Our findings point to urgent gaps in water quality management that must be addressed to reduce exposure to *B. pseudomallei* and other waterborne pathogens.

We have reported these findings to the Thailand Provincial Waterworks Authority and the Regional Ministry of Public Health to support immediate policy review. In line with Sustainable Development Goal 6—ensuring access to clean water and sanitation for all—our results underscore that improving water treatment, monitoring, and infrastructure maintenance can substantially reduce the risk of melioidosis. Targeted public health interventions and investment in water safety are essential to protect vulnerable communities and lower disease burden.

## Materials and methods

### Study design

We conducted two inter-related studies to improve *B. pseudomallei* detection sensitivity in environmental samples and to assess the association between environmental presence and melioidosis clinical incidence.

**Study 1** evaluated the performance of CRISPR-BEEPs against a conventional approach, with two qPCR assays as the reference standard. A total of 356 water samples were collected between November 2020 and November 2021 from households and natural water sources used by 243 participants, who were part of the case-control cohort from **Study 2** in Ubon Ratchathani and nearby provinces in northeast Thailand, areas known to be endemic for melioidosis. This study adhered to the Standards for Reporting of Diagnostic Accuracy (STARD) guideline.

**Study 2** investigated the association between the presence of *B. pseudomallei* in water samples and the incidence of melioidosis in households of cases and controls. From October 2019 to January 2023, a total of 1,135 participants were recruited from a melioidosis cohort. This cohort consisted of 439 melioidosis patients, 190 patients with other community-acquired infections, and 506 healthy controls. Melioidosis and other infectious cases were recruited immediately after a culture-confirmed diagnosis at Sunpasitthiprasong Hospital, using hospital computer records. Controls were selected from blood donors or diabetic outpatients at the same hospital. After obtaining informed consent, demographic information such as age, sex, ethnicity and underlying health conditions was extracted from medical records, while participants provided details about their exposure risks and household locations (**Supplementary methods**).

The participants whose water samples were collected in Study 1 included 70 melioidosis patients, 69 patients with other bacterial infections, and 104 healthy controls from Study 2. To address sample variability, multiple water samples were collected from the household and surrounding areas of 80 participants, including 74 cross-seasonal samples to account for seasonal fluctuation (**Supplementary Figure 3**). This study received ethical approval from the Sunpasitthiprasong Hospital Ethical Review Board (015/62C) and the Oxford Tropical Research Ethics Committee (OxTREC 25-19). The full study protocol is described in ^49^ and followed the Strengthening the Reporting of Observational Studies in Epidemiology (STROBE) guideline.

### Environmental sample collection and enrichment

Five-litre water samples were collected from each household, except during the peak of SARS-CoV-2 outbreaks, when nearby water reservoirs were sampled due to visitation restrictions. Participants’ water sources included pipe water, boreholes, ponds, lakes, canals or rivers (**Figure 3c**, **Supplementary Figure 1**). Samples were transported in sterile plastic bags to the laboratory within three hours and processed promptly. Each sample was filtered (0.45-μm cellulose, Sartorius, Germany), and cultured on Ashdown^65^ agar to enrich for *B. pseudomallei* growth. Plates were incubated at 40 °C, and inspected daily for four days; plates with fungal overgrowth were discarded.

### Conventional and alternative screening

For conventional screening, colonies showing typical *B. pseudomallei* morphologies were confirmed using monoclonal antibody-based assays^58,59^ (**Supplementary Figure 2**). For the CRISPR-BEEPs and qPCR tests, microbial lawn, including both visible and non-visible *B. pseudomallei,* were collected from the plates and preserved in glycerol stock (20% v/v, VWR, Belgium) at -80 °C for subsequent DNA extraction. All procedures were conducted in an enhanced biosafety level 2 laboratory but with biosafety level 3 practices. For each type of screening, the personnel performing the tests were blinded to the results of the other methods.

### Bacterial DNA extraction

Microbes were revived from plate-sweep glycerol stocks for each sample. A loop of colonies was used for bacterial DNA extraction with QIAGEN Genomic-tips [Cat#10243, Germany] to preserve high molecular-weight pan-microbial DNA and minimise DNA shearing (**Supplementary method**). The extracted DNA had an average concentration of 278.42 ng/μL (IQR 181.00 – 433.23) and an A260/A280 purity score ranging from 1.68 and 1.94, ensuring high yield and low impurities, suitable for CRISPR-BEEPs and qPCR detection.

### *In silico* evaluation of qPCR (*TSS1*, *BPSS1386*) and CRISPR-BEEPs (crBP34 + RPA) target specificity and coverage

To evaluate the specificity of the selected DNA targets, we performed *in silico* screening using BLASTn^66^ (v.2.16.0) against 27,771 metagenome-assembled genomes (MAGs) derived from ecological samples^41^ collected across tropical and subtropical regions. The screened sequences include the full amplicons generated by the forward and reverse primers of the *TSS1* (115 bp), *BPSS1386* (185 bp), and CRISPR-BEEPs which encompassing the RPA primer sites and *crBP34* region (118 bp) (**Supplementary Table 1, Supplementary Figure 5**). All partial hits were examined for mismatches and their locations, as the position of mismatches influences assay performance. Based on published tolerance thresholds, we considered up to three mismatches outside the 3’ end for qPCR primers^44,45^, up to three non-clustered mismatches and outside 3’ end for RPA primers^51^, and one to two mismatches in non-seed regions for the CRISPR-Cas12a crRNA^52^ as acceptable.

To assess the conservation of these targets across the global *B. pseudomallei* population, we also screened the same amplicons to 3,341 high-quality *B. pseudomallei* genome assemblies^46–50^ using BLAT (v. 36) as described in ^38^. Partial matches were evaluated using the same mismatch criteria as described above.

### qPCR detection and determination of *B. pseudomallei* positive environmental samples

Two independent qPCR primer sets - *TTS1* and *BPSS1386* - were used to cross-validate each other’s results. Primers were tested for efficiency (**Supplementary Figure 5**). Each DNA sample was tested in duplicate with both primer sets. Reactions were performed in a 20-μL master mix volume. The protocol included an initial denaturation step at 95 C for 10 minutes, followed by 40 cycles of denaturation at 95 C at 15 seconds, with annealing temperature at 61 C for *TTS1* and 64 C for *BPSS1386* primers. Samples were processed in batches alongside known positive and negative controls. The cycle threshold (Ct) values were recorded for each sample. The individuals interpreting the results were blinded to the source of water samples. A sample was classified as positive if qPCR amplification occurred for both primers and the melting temperatures were consistent with the expected values. Samples were classified as inconclusive if amplification with correct melting temperature was observed with only one primer. Samples were considered negative if no amplification was detected or if amplification occurred with incorrect melting temperatures.

### CRISPR-BEEPs detection

The extracted DNA samples were amplified using the Recombinase Polymerase Amplification (RPA) method with TwistAmp® Basic (Cat#TABAS03KIT, TwistDx, Maidenhead, UK). RPA primers (**Supplementary Table 1**) were used at a final concentration of 0·48 nM, and 2 - 6 μL of genomic DNA were added to a 30-μLRPA reaction, which was incubated at 39 °C for 30 minutes. The resulting amplicons were added to CRISPR reaction (**Supplementary method**) and incubated at 37 °C for 60 minutes. A HybriDetect universal lateral flow assay kit (Cat#MGHD1, Milenia Biotec, Giessen, Germany) was dipped into the CRISPR reaction for 5 minutes. Result from the lateral flow kit was observed visually by three individuals, with interpretations made independently and without reference to the outcomes of other tests. Two bands indicated a positive result, while a single band signified a negative result.

### Determination of molecular sensitivity for qPCR and CRISPR-BEEPs

We conducted a spiking experiment where we introduced varying concentrations of *B. pseudomallei* DNA (0, 2.5, 5, 10, 20, 50, and 250 copies/μL) into a 2 μLvolume of pan-microbial DNA background (Figure 2c). This background DNA was derived from common bacterial species^53^ identified in water sources including *Escherichia coli, Klebsiella pneumoniae, Enterobacter cloacae, Serratia odorifera, Citrobacter freundii,* and *Leclercia adecarboxylata.* The pan-microbial DNA was prepared at a final concentration of 278 ng/µL, representative of the range observed in our actual samples. This setup mimicked the mixed microbial environment of the water samples and enabled us to evaluate the minimum concentration of *B. pseudomallei* detectable by both the qPCR and CRISPR-BEEPs methods.

### Statistical analysis

For study 1, the minimum sample size was calculated using the formula n = z^2^ x p x (1-p)/d^2^, with “z” as the 95% confidence interval at 1.96; “p” representing the prevalence at 0.5; and “d” as the margin of error at 0.1. At least 96 positive and 96 negative environmental samples were needed to validate the conventional approach and CRISPR-BEEPs. Sensitivity and specificity were computed using double-qPCR results as the reference, with the 95% confidence interval estimated based on a binomial assumption. McNemar’s test was used to compare detection performance with paired data.

For study 2, the proportion of *B. pseudomallei* positive samples per area was calculated for each method (double-qPCR, CRISPR-BEEPs, and conventional) by determining the number of positive samples relative to the total collected within the radii of 1 km to 10 km from participant households (**Supplementary Tables 4-13**). This measure is referred to as the positivity rate in neighbourhood water reservoirs. Distances were computed based on each participant’s geoposition using the R package “geosphere”^67^. The association between *B. pseudomallei* in household and neighbourhood water reservoirs, participants’ occupations, health conditions, and disease status were analysed using univariable (**Supplementary Table 3**) and multivariable logistic regression models^68^ (**Figure 5e**, **Supplementary Tables 4-13**). All tests were two-sided with a significance level of 0.05, and analyses were conducted using R (version 4.3.2), including data visualisation.

Sensitivity analyses were conducted to evaluate the robustness of the results under various conditions. First, we restricted the study population to the actual water sampling period from November 2020 to November 2021. Second, we expanded the study period to include data from October 2019 to January 2023. Third, we assessed the geographical ranges of exposure by using different radius distances from each household. Additionally, we analysed the data by season to account for observed bacterial persistence in the environment across different seasons and the potential latency period, which may span several years before melioidosis develops^7^.

## Data Availability

All data produced in the present study are available upon reasonable request to the authors

## Contributors

SP, CCho and PB contributed equally. CChe, NPJD and NRT conceived the cohort study. CChe and JCo conceived the additional water sample activities. CChe, SPW and CU conceived the CRISPR-based study. CChe and SP secured funding. CChe, SPW, CU, NRT, JP, NPJD, SJP, PC and SS designed the study. SP, CCho, PB, KA, and CChe collected water samples. SP, CCho, PB, YD, AF, GW, and PA processed water samples. GW, PA, and VW provided conventional detection methods. SPW and CU provided molecular detection methods. JP and NRT provided *in silico* detection methods. SP, KA, AW prepared data for the analysis. CChe did the analysis. SP, CCho, PB, AW, SJP, JCh, JCo, JP, CU, SPW, CChe interpreted the data. JCh communicated with Thailand Provincial Waterwork Authorities and proposed engineering resolutions to reduce risks. SP, CCho, PB, AW, and CChe assessed and verified underlying data of the study. All authors had full access to all data in the study and had final responsibility for the decision to submit for publication. CChe wrote the first and revised draft. All authors read and approved the manuscript.

## Data availability

The genomic data on microbial communities and bacterial genomes used in this study are publicly available^41,46–48,50^. De-identified participant data supporting the findings of this article are available upon request. Researchers should contact the corresponding author at. All requests will be reviewed to ensure compliance with informed consent and intended use for research purposes. Rain and flood data in Ubon Ratchathani are publicly available from Thailand Royal Irrigation Department (https://www.rid.go.th/index.php/th/).

## Acknowledgements

CChe was funded by the Wellcome International Intermediate Fellowship (216457/Z/19/Z), the Sanger International Fellowship, and the University of Oxford Nuffield Department of Medicine Career Development Scheme. This research was funded in part by the Wellcome Trust [220211 and 206194] and Thailand Science Research and Innovation, Fundamental Fund 2024 [FRB670026/0457, to CU]. For the purpose of Open Access, the author has applied a CC BY public copyright license to any Author Accepted Manuscript version arising from this submission. The funders had no role in study design, data collection and analysis, decision to publish, or preparation of the manuscript.

