## Supplementary Information for "CRISPR-based environmental detection of *Burkholderia pseudomallei* identifies sanitation gaps and melioidosis risk in northeast Thailand"

**This Supplementary Information includes Supplementary Methods, Supplementary Figures 1-5, Supplementary Table 1 – 13, and Supplementary References**

### Supplementary methods

#### STUDY 1 PROTOCOL: ENVIRONMENTAL SCREENING FOR *B. PSEUDOMALLEI*

This study aims to compare and improve the environmental screening protocols for *Burkholderia pseudomallei* by comparing the conventional approach[1] against the alternative CRISPR-BEEPS method[2,3], using two primer-based qPCR assays as reference tests.

#### Reviews of existing methods of water sample surveillance for *B. pseudomallei*

The current guideline[1] for environmental sampling of *B. pseudomallei* was established in 2013 by the Detection of Environmental *B. pseudomallei* Working Party (DEBWorP). This guideline focuses on soil and water sampling, primarily using conventional culture-based methods on selective media, followed by visual inspection of colonies that resemble *B. pseudomallei*. These traditional methods have been widely used to study the environmental presence of *B. pseudomallei*, helping define its ecological habitats in regions such as Australia, Oceania, Southeast Asia, Africa, and South America [4–12]. For water sampling, the protocol involves collecting 1 to 5,000 mL of water and concentrating the bacteria using filtration, centrifugation, or potassium alum precipitation. Detection follows selective media, such as Ashdown[13] agar containing gentamicin and crystal violet, after which colonies are visually examined for characteristic *B. pseudomallei* morphologies and isolated for subsequent characterisation. However, the rise of antibiotic-resistant microbes complicates the differentiation of *B. pseudomallei* from other environmental bacteria.

To improve detection, recent studies have introduced molecular methods such as PCR-based assays[14–17], mass spectrometry[18,19], and sequencing[19,20], often following culture with Ashdown agar or modified Ashdown broth. qPCR-based methods have shown a significant increase in test positivity rate compared to conventional using Ashdown agar alone. In Laos, qPCR nearly doubled the positivity rate when applied to the same water samples[21]. Similarly, a study in Australia demonstrated that a qPCR method using samples enriched in Ashdown's broth achieved a positivity rate of 69.2%, while the conventional method yielded only 5.5%[22]. Although effective, molecular techniques often require complex and costly equipment, which may not be available in resource-limited settings. To overcome this issue, we have developed CRISPR-BEEPS[2,3], an assay that detects *B. pseudomallei* nucleic acids without complex equipment, making it suitable for resource-limited areas.

#### Water collection and enrichment

In summary, five liters of water were collected from either household tap water or a neighborhood water reservoir[23] (**Supplementary Figure 1**). For this study, one liter was used, which was passed through two 0.45-μm filters, with 500 mL filtered through each. Both filters were then cultured on Ashdown agar to isolate *B. pseudomallei* for further confirmation.

#### Conventional plate inspection method

The conventional method, including confirmation using monoclonal antibodies (**Supplementary Figure 2**), was carried out following the procedures outlined in [24].

#### An alternative molecular method

##### *Plate-sweep approach*

The Plate Sweep method begins with recovering *B. pseudomallei* or other Gram-negative bacteria from lawn mixed colonies on Ashdown agar (**Supplementary Figure 2**). The bacteria colonies are then harvested by sweeping the plate with a 10 µL-loop, after which the bacterial pellet is resuspended in 5 mL of PBS to wash the cells. It is important to ensure all representative strains are included by sweeping from the first plate. The mixture is centrifuged at 3000g for 5 minutes, and the supernatant is removed by quickly decanting the liquid into a waste bin. The bacterial pellet is resuspended in 5 mL of PBS, vortexed at top speed to ensure a homogeneous mixture, and centrifuged again at 3000g for 5 minutes before the supernatant is removed. All procedures were conducted in an enhanced biosafety level 2 laboratory but with biosafety level 3 practices.

##### *DNA extraction method used in this study*

For DNA extraction, 7 µL of RNase A solution (100 mg/mL) is added to 3.5 mL of Buffer B1, which should be prepared fresh and stored at 2-8°C. Lysozyme is dissolved in distilled water to a concentration of 100 mg/mL and stored at -20°C, while Buffer QF is warmed to 50°C to increase the DNA yield. Isopropanol and 70% v/v ethanol should also be stored at -20°C and placed on ice during the procedure.

Bacterial lysis begins by resuspending the bacterial pellet in 3.5 mL of Buffer B1 (with RNase A) and vortexing it thoroughly. Then, 80 µL of lysozyme stock solution (100 mg/mL) and 100 µL of proteinase K are added, followed by incubation at 37°C for at least 30 minutes. Next, 1.2 mL of Buffer B2 is added, mixed by inverting the tube several times, and incubated at 50°C for 1-2 hours until the mixture becomes clear, indicating complete lysis. If lysis is incomplete, extend the incubation or vortex the mixture before separating the bacterial debris by centrifugation at 5000g for 10 minutes at 4°C.

For DNA purification, a Qiagen genomic tip is equilibrated with 4 mL of Buffer QBT when 10 minutes remain in the previous incubation. The lysed sample is then applied to the genomic tip using a Pasteur pipette and allowed to pass through by gravity flow. The genomic tip is washed twice with 7.5 mL of Buffer QC.

During DNA elution and precipitation, the genomic DNA is eluted with 5 mL of prewarmed Buffer QF and collected in a new 50 mL centrifuge tube. Depending on the volume, 800 µL of the eluted DNA is aliquoted into labelled 2 mL microcentrifuge tubes. DNA precipitation is achieved by adding 560 µL of ice-cold isopropanol and inverting the tubes 20-30 times until the DNA precipitates as a white, feather-like strand. If the DNA is not visible, proceed with the steps regardless. The DNA is stored at -20°C overnight.

The following day, the tubes are kept on ice, and the precipitated DNA is centrifuged at maximum speed at 4°C for 5 minutes. The supernatant is carefully discarded, and the DNA pellet is washed with 1 mL of ice-cold 70% v/v ethanol. An alternative method involves transferring the DNA pellet into one tube with 200 µL of ethanol, creating a final volume of 1.2 mL. The DNA is centrifuged again, and the remaining supernatant is removed carefully using a P20 pipette if necessary. The DNA pellet is air-dried for 5-10 minutes, ensuring it does not overdry. If large droplets remain, incubate the pellet on a 55°C heat block for 3-5 minutes.

To dissolve the DNA, add 35 µL of nuclease-free water (or ~200 µL for the alternative method) and incubate it overnight or at 55°C for 1-2 hours. Finally, the DNA is stored at 2-8°C for short-term storage or at -20°C for up to 6 months.

**CRISPR-BEEPs**

***Expression and purification of MBP-LbCas12a***

The expression and purification of MBP-LbCas12a were performed according to the procedures detailed in the previous study[2].

***CRISPR RNA Synthesis***

The CRISPR RNA, crBP34, previously reported[2], was used in this study. The synthesis of crRNA followed the same methods outlined in the prior study.

***Recombinase polymerase amplification (RPA)***

RPA was performed using the TwistAmp Basic kit (TwistDx, USA, #TABAS03KIT) following the manufacturer's protocol, with the following modifications:
(i) The total reaction volume was adjusted to 30 µL,
(ii) Incubation was carried out at 39°C for 30 minutes,
(iii) DNA input was limited to 2 µL, as adding more DNA did not improve sensitivity and could introduce inhibitors, and
(iv) MgOAc was added last to initiate the reaction. All RPA reactions were stored at -20°C until use.

RPA primers 148 and 149 (**Table S2**), previously reported[2], were synthesised and purified by Macrogen (Korea) using standard desalting.

***CRISPR reaction and dipstick detection***

CRISPR reactions were conducted in a 50-µL volume containing 100 nM crRNA, 200 nM MBP-LbCas12a, 100 nM FAM-biotin probe, 5 µL RPA product, and 1x HOLMES buffer (2 mM spermidine, 40 mM Tris pH 8.5, 6 mM $MgCl_2$, 1 mM DTT, 40 mM glycine, 0.001% v/v Triton X-100, and 0.4% w/v PEG-20,000). The reaction was incubated at 37°C for 60 minutes. After incubation, a lateral flow dipstick (Milenia Biotec, Germany, #MGHD1) was inserted into the reaction mixture and allowed to develop for 5 minutes before reading the results.

***qPCR***
Two sets of qPCR primers, TTS1 and BPSS1386 (**Table S2**), were used to detect *B. pseudomallei* genomic DNA in samples. Since environmental microbes can exchange genetic material via horizontal gene transfer, both primer sets, targeting different genes, were employed to ensure the accuracy of the detection. The primers were synthesised and desalted by Macrogen (South Korea).

qPCR reactions were prepared in a 20-µL volume using Maxima SYBR Green/ROX qPCR Master Mix (ThermoFisher Scientific, USA, #K0221), following the manufacturer's protocol. The qPCR conditions included: an initial denaturation at 95°C for 10 minutes, followed by 40 cycles of denaturation at 95°C for 15 seconds, with annealing temperatures of 61°C for TTS1 and 64°C for BPSS1386. Each sample was processed in duplicate along with known positive and negative controls. The cycle threshold (Ct) values were recorded for each sample, and positive detection was defined as any duplicate reaction yielding a positive result for either primer set. Standard melting curve analysis was performed at the end of the PCR and compared to the melting profiles of the positive controls. The ΔRn threshold was automatically determined by the built-in software.

### STUDY 2 PROTOCOL: A CASE-CONTROL OBSERVATIONAL COHORT STUDY

#### Study design, sites and population

One of the objectives of this study was to investigate the risk factors associated with melioidosis infection. To achieve this, demographic information, occupation, and environmental exposure data, along with water samples were collected from participants, including melioidosis cases, other infections, and healthy controls, living within a 120 km radius of Sunpasitthiprasong Hospital in Ubon Ratchathani and nearby provinces, where high incidences of melioidosis are observed. The hospital, which hosts a satellite unit of the Mahidol-Oxford Tropical Medicine Research Unit (MORU), also facilitated the collection and processing of water samples for the study. The full study protocol is available in Angchagun *et al.* 2023[23].

Care was taken to maintain the separation of cases and controls throughout the study. The control group had no documented history of melioidosis at the point of the study end date in January 2023. We acknowledged the transfer of controls to cases. Throughout the study period, one control with underlying diabetes has become infected with melioidosis and thus was transferred into the case category. This accounted for 0.2 % of the control population.

Given the similar clinical manifestations between melioidosis and tuberculosis (TB), individuals with active TB were excluded from the study. Additionally, other underlying health conditions that may increase susceptibility to bacterial infections were also excluded. The complete inclusion and exclusion criteria are outlined below.

#### Participant inclusion criteria

**Melioidosis cases:**

- Participants must be aged 18 years or older.

- A culture-confirmed infection of *B. pseudomallei* must be obtained from any clinical sample.
- Participants must be willing to participate in the study, with written informed consent provided by the patient or their relative.
- Participants must have resided in northeast Thailand for at least the previous two years.

**Other Infection controls:**
- Participants must be aged 18 years or older.
- A culture-confirmed infection by bacterial pathogens other than *B. pseudomallei* must be obtained.
- Participants must be willing to participate in the study, with written informed consent provided by the patient or their relative.
- Participants must have resided in northeast Thailand for at least the previous two years.

**Healthy controls:**
- Participants must be aged 18 years or older.
- Participants should currently be in good health (non-diabetic) or be outpatients from a diabetic clinic, without any other medical problems requiring hospital supervision.
- Participants must be willing to participate in the study, with written informed consent provided by the patient or their relative.
- Participants must have resided in northeast Thailand for at least the previous two years.

**Participant exclusion criteria**
**Melioidosis cases:**
- Current tuberculosis (TB) or treatment for TB within the last six months.
- Documented human immunodeficiency virus (HIV) infection, chemotherapy, or other immunosuppressive therapies in the last 12 months.
- Pregnancy.

**Other Infection controls:**
- Previous history of melioidosis.
- Current tuberculosis (TB) or treatment for TB within the last six months.
- Documented human immunodeficiency virus (HIV) infection, chemotherapy, or other immunosuppressive therapies in the last 12 months.
- Pregnancy.

**Healthy controls:**
- Previous history of melioidosis.
- Current tuberculosis (TB) or treatment for TB within the last six months.
- Documented human immunodeficiency virus (HIV) infection, chemotherapy, or other immunosuppressive therapies in the last 12 months.
- Pregnancy.

**Ethics/protection of human subjects**

**Screening**

All participants meeting the recruitment criteria were approached by a research nurse for eligibility screening. A written participant information sheet (PIS) and informed consent form (ICF) in Thai were provided to participants or their relatives if they lacked capacity. The PIS detailed the study's nature, implications, constraints, known side effects, and risks. Participants were given a minimum of ten minutes to consider the information and had the opportunity to ask questions before deciding whether to participate.

Informed consent was obtained through a dated signature from the participant or their relative, as well as from the person obtaining consent. For illiterate participants, a thumbprint was used. A signed copy of the ICF was provided to the participant or responsible relative, while the original signed forms were retained at the study site.

**Ethics/Protection of Human Subjects**

The study received ethical approval from the Sunpasitthiprasong Hospital Ethical Review Board (015/62C) and the Oxford Tropical Research Ethics Committee (OxTREC 25-19). All research data sets were pseudonymised to protect participant identities and allow for feedback or withdrawal of consent. No participant names, identities, or exact GPS locations of their households will be disclosed.

**Other prevalent bacterial infections identified in other infectious controls**

Among the infection control group (n = 190), the most prevalent infections were *Escherichia coli* (n = 65, 35%), followed by *Klebsiella pneumoniae* (n = 40, 21%), coagulase-negative *Staphylococcus spp.* (n = 11, 5.9%), and *Acinobacter baumanii* (n = 9, 5.3%). One patient was co-infected with both *K. pneumoniae* and *A. baumanii.* These findings align with previous studies on community-acquired infections in adults from northeast Thailand (2013-2017[25] and 2004-2010[26]) and a broader study in rural communities in Southeast Asia[27].

**Examples of cases and controls with multiple water samples collected**

**Melioidosis case**

A monk reported using mixed water sources, including piped water and water pumped from nearby ponds. His daily alms round, a mindfulness practice where he walked barefoot through the village to receive food from villagers, further exposed him to potential environmental risks. Five water samples were taken: from the temple's piped water, the pond water used in the temple, a pond at the temple entrance, a canal flowing past the temple and village, and a lake serving as a water reservoir. All but the pond at the temple entrance tested positive for *B. pseudomallei* (4 out of 5 samples, 80%). The monk had been admitted for melioidosis earlier that year.

**Healthy control**

A homemaker reported using public piped water at home. Five water samples were collected, including household piped water and ponds surrounding her village. Three pond water samples tested positive for *B. pseudomallei* (3 out of 5 samples, 60%). Although her daily routine was home-based, making environmental exposure unclear, she reported not having melioidosis up to the time of enrollment.

**Other infection control**

A third case is an employer who used public piped water at home. Five water samples were collected, including a sample from household piped water and four samples from nearby ponds. Only one pond sample tested positive for *B. pseudomallei* (1 out of 5 samples, 20%). He was infected with *Escherichia coli* and had no reported history of melioidosis at the time of the study.

**Supplementary Figures**

**Supplementary Figure 1 Type of water samples collected**

This included piped water, surface water from non-groundwater reservoirs such as rivers, canals, lakes, and ponds, and groundwater sourced from boreholes. These samples represent the potential environmental exposures for individuals living in the study areas.

**Supplementary Figure 1**

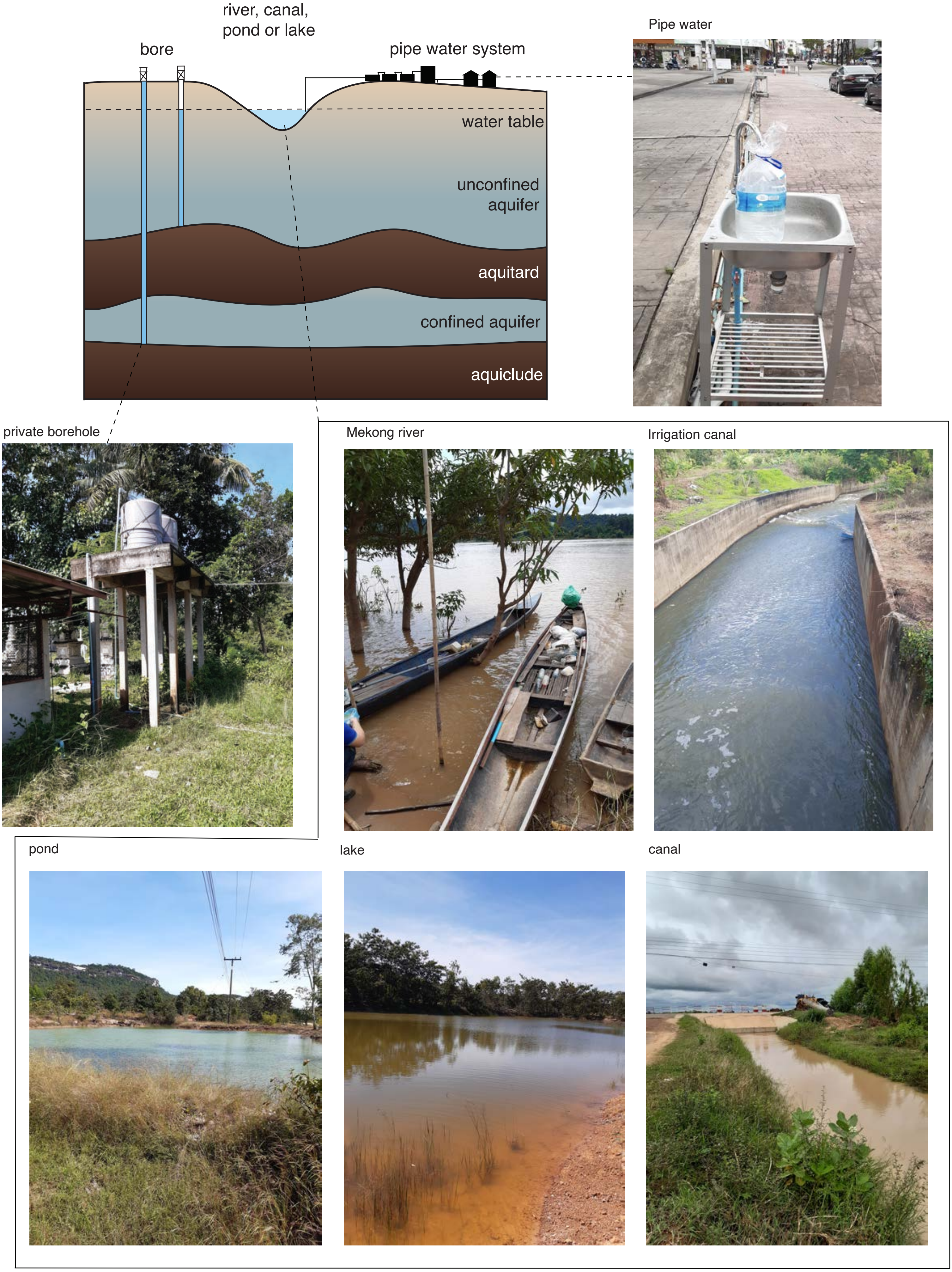

**Supplementary Figure 2 Screening approaches**
(a) Morphological characteristics of *B. pseudomallei* used for species identification on Ashdown agar plates. (b) Conventional method: A sample of suspected *B. pseudomallei* grown on filtered water paper is placed on an Ashdown agar plate. Suspected colonies (white circles) were re-cultured onto fresh Ashdown agar and confirmed using monoclonal antibody tests. (c) Alternative plate-sweep approach: bacteria grown in modified Ashdown broth[21,22] or on filtered papers placed on Ashdown agar were plate-swept for DNA extraction, followed by testing with CRISPR-BEEPs (the assay used in this study) or PCR (reference assay).

**Supplementary Figure 2** Typical morphology of *Burkholderia pseudomallei* and comparison between conventional and alternative plate-sweep screening methods for water samples

**a Morphology of *Burkholderia pseudomallei* colonies on solid media like Ashdown's agar**

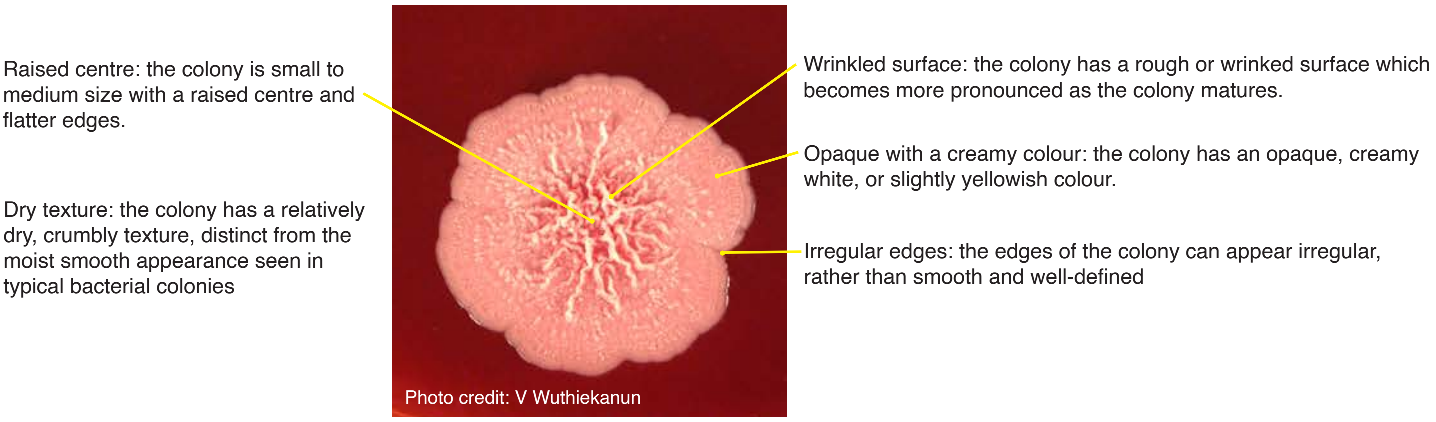

**b Conventional plate inspection followed by species confirmation with monoclonal antibodies**

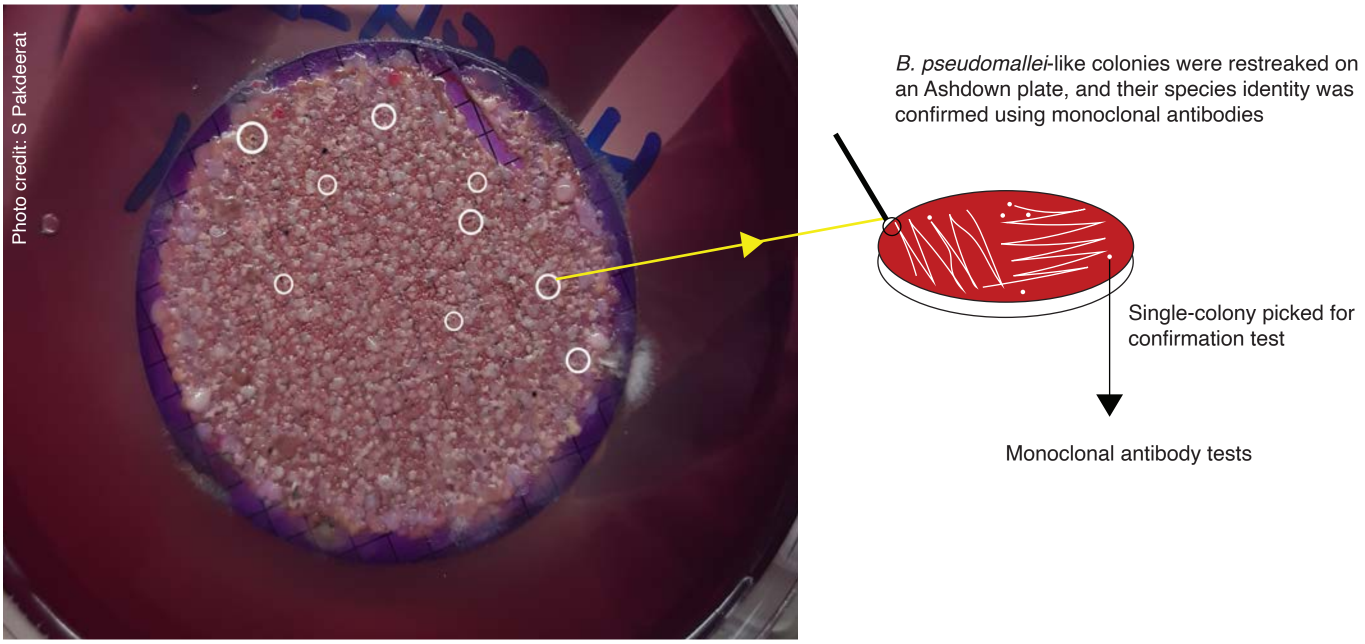

**c New plate-sweep method followed by species confirmation with PCR or CRISPR-BEEPs**

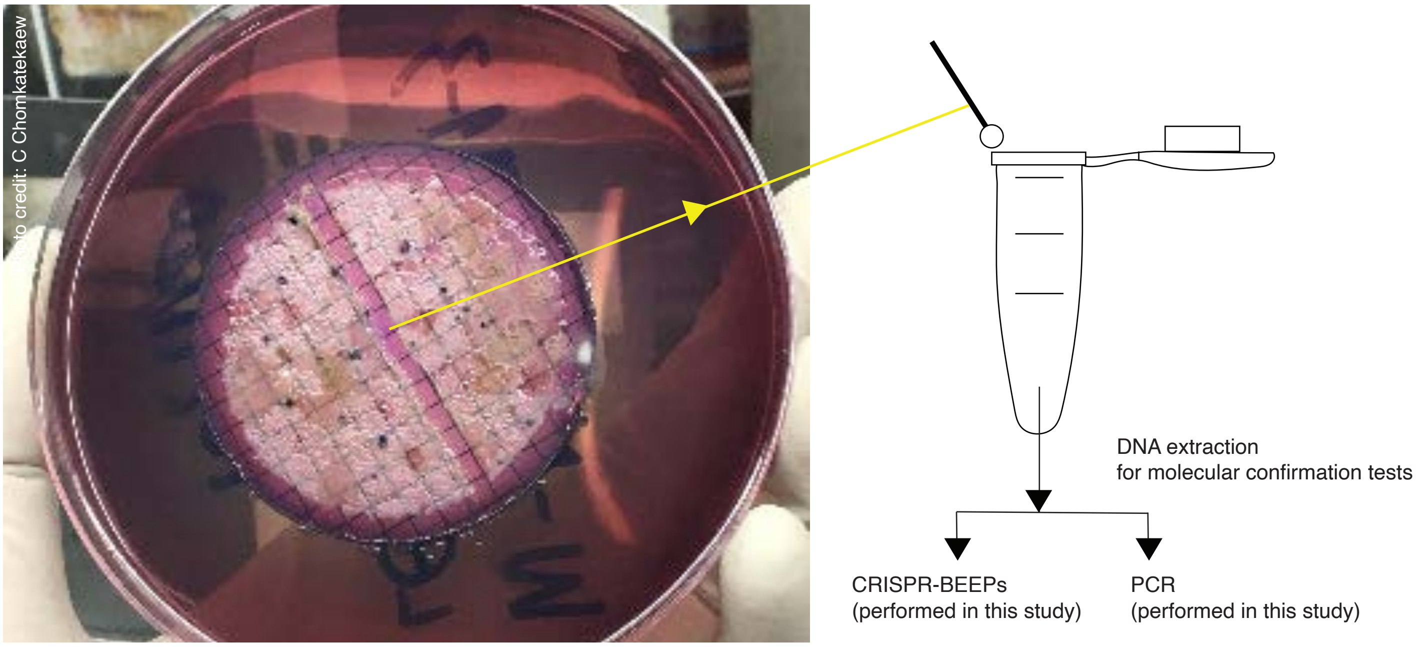

**Supplementary Figure 3 Participants with multiple water samples collected.**
(a) Temporal distribution of collected water samples by participant group, presented from left to right: healthy controls, other infectious controls, and melioidosis cases. Each row represents an individual participant, with dots indicating the collected water samples; the size of each dot is proportional to the sample size. (b) *B. pseudomallei* positivity rates in water samples collected from participants across different seasons, categorised by case and control groups. The upper arc represents the dry season, while the lower arc indicates the rainy/flood season. In both (a) and (b), colour coding reflects the positivity of *B. pseudomallei* in water samples, as determined by double-primer qPCR assays.

**Supplementary Figure 3**

**a** participants with multiple samples collected and the seasonal fluctuation in positive *B. pseudomallei* samples

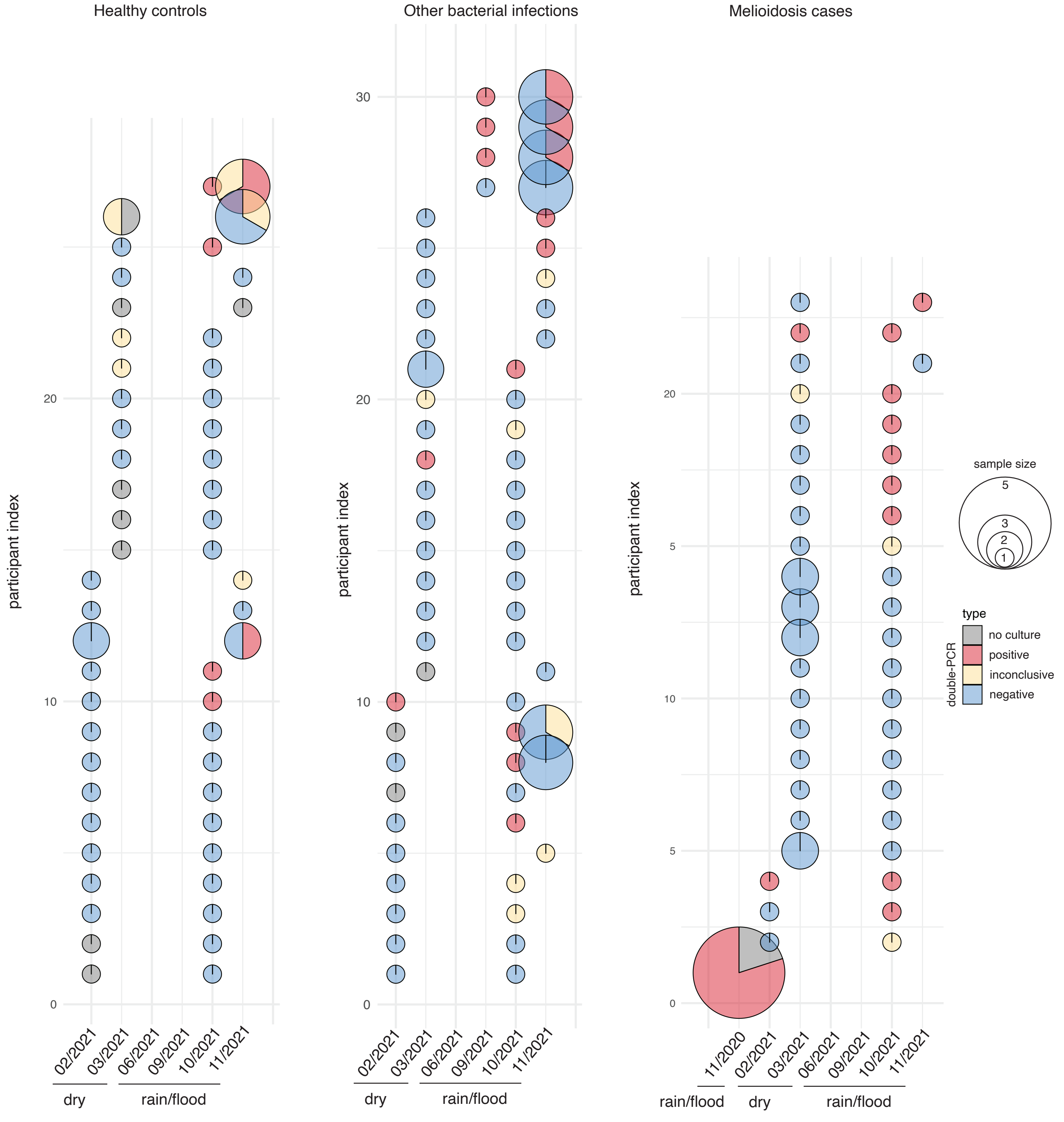

**b** Changes in *B. pseudomallei* positivity rate between the dry and rainy/flood seasons across cohort

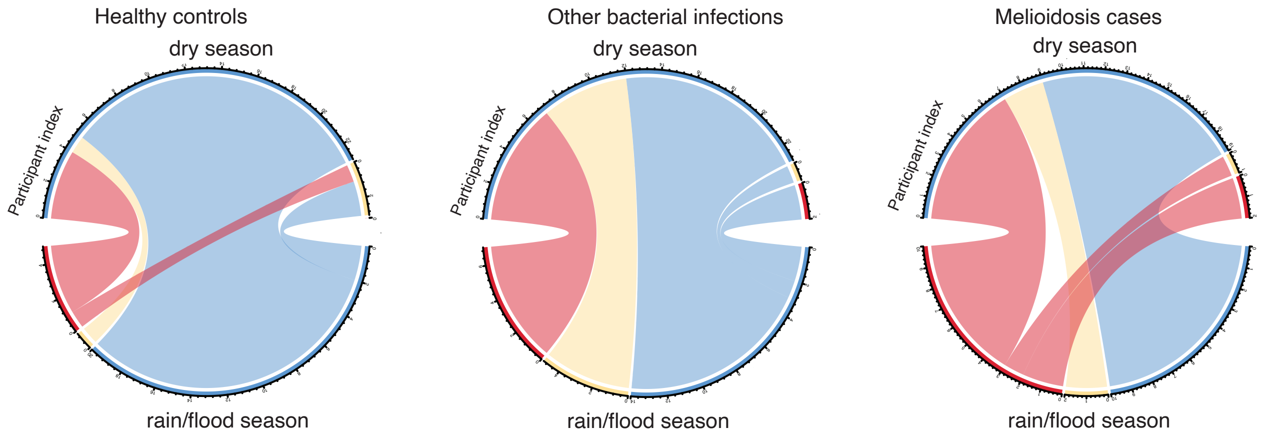

**Supplementary Figure 4 *B. pseudomallei* positivity rates in water samples**

(a) Positivity rates in water samples collected directly from households (n = 243) were compared across household types using two methods. First, a whole count comparison was conducted where any household with at least one positive sample was classified as positive, and differences between positive and non-positive households were analysed using the two-tailed Fisher's exact test. Second, positive rates were calculated for households with multiple samples by determining the rate of positive samples per household. These positive rates were then compared across household types using the two-tailed Wilcoxon signed-rank test. (b) For positivity rates in water samples collected indirectly from collection points within a 1 to 10 km radius of the households, the analysis also included households that did not have direct water samples but were located near collection points. As part of a sensitivity analysis, positivity rates were calculated based on water samples collected within various distance (1 to 10 km) from each participant's household. Differences in positivity rates across household categories were assessed using the non-parametric Wilcoxon test.

**Supplementary Figure 4**

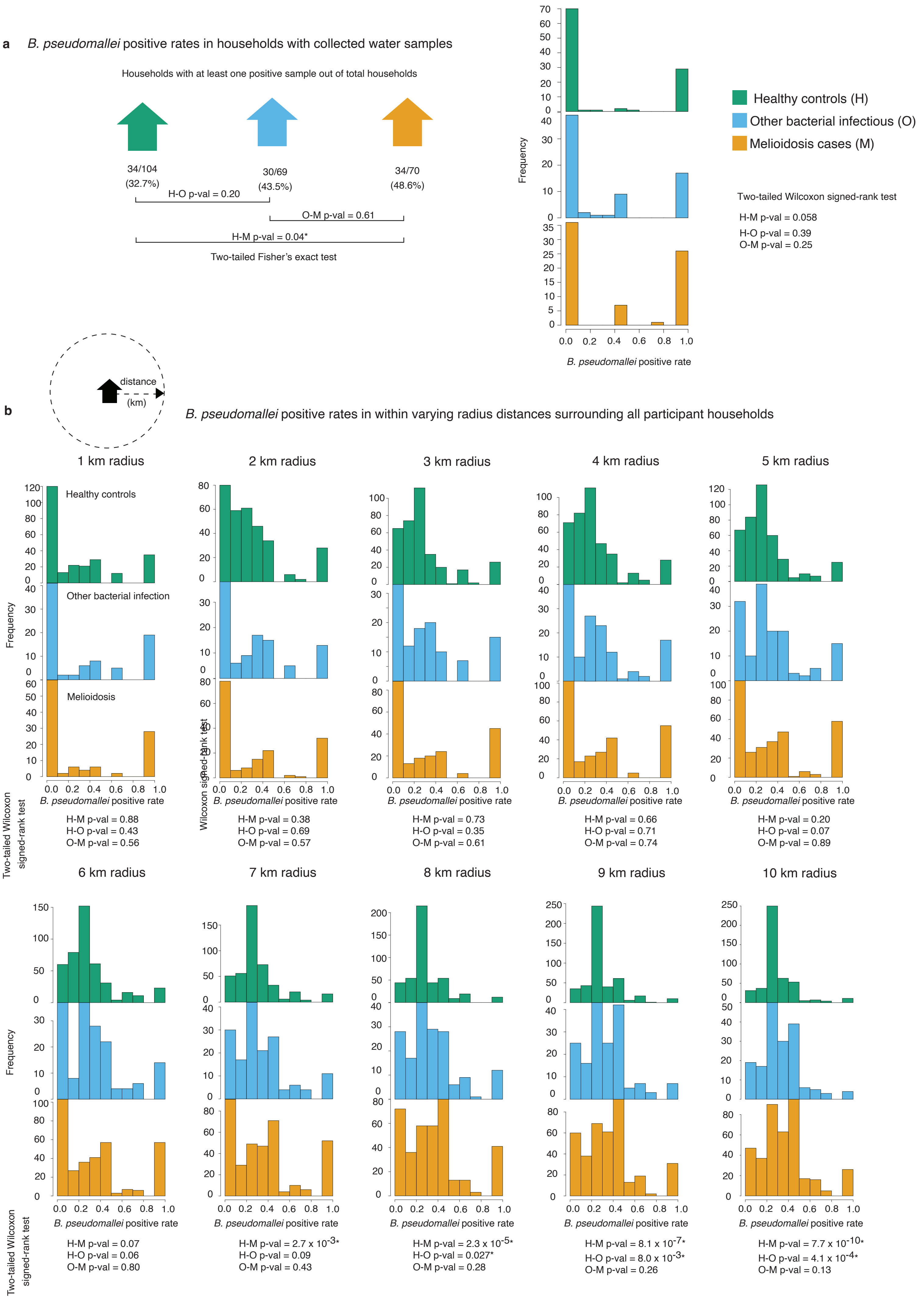

**Supplementary Figure 5 Efficiency, coverage, cycle threshold (ct) scores of PCR primers used in this study.**

(a) *In silico* PCR analysis demonstrating the detailed coverage of primer 1 (*TTS1*) and primer 2 (*BPSS1386*) across the *B. pseudomallei* genomic population. (b) Linear regression analysis used to calculate primer efficiencies: *TTS1* (96.1%) and *BPSS1386* (81.3%), determined using a formula in reference[28]. (c) Ct values for each experimental replicate using primers *TTS1* and *BPSS1386*. Black points indicate experiments with correct melting temperatures, while grey points represent those with incorrect melting temperatures, where non-specific primer binding may occur.

**Supplementary Figure 5**

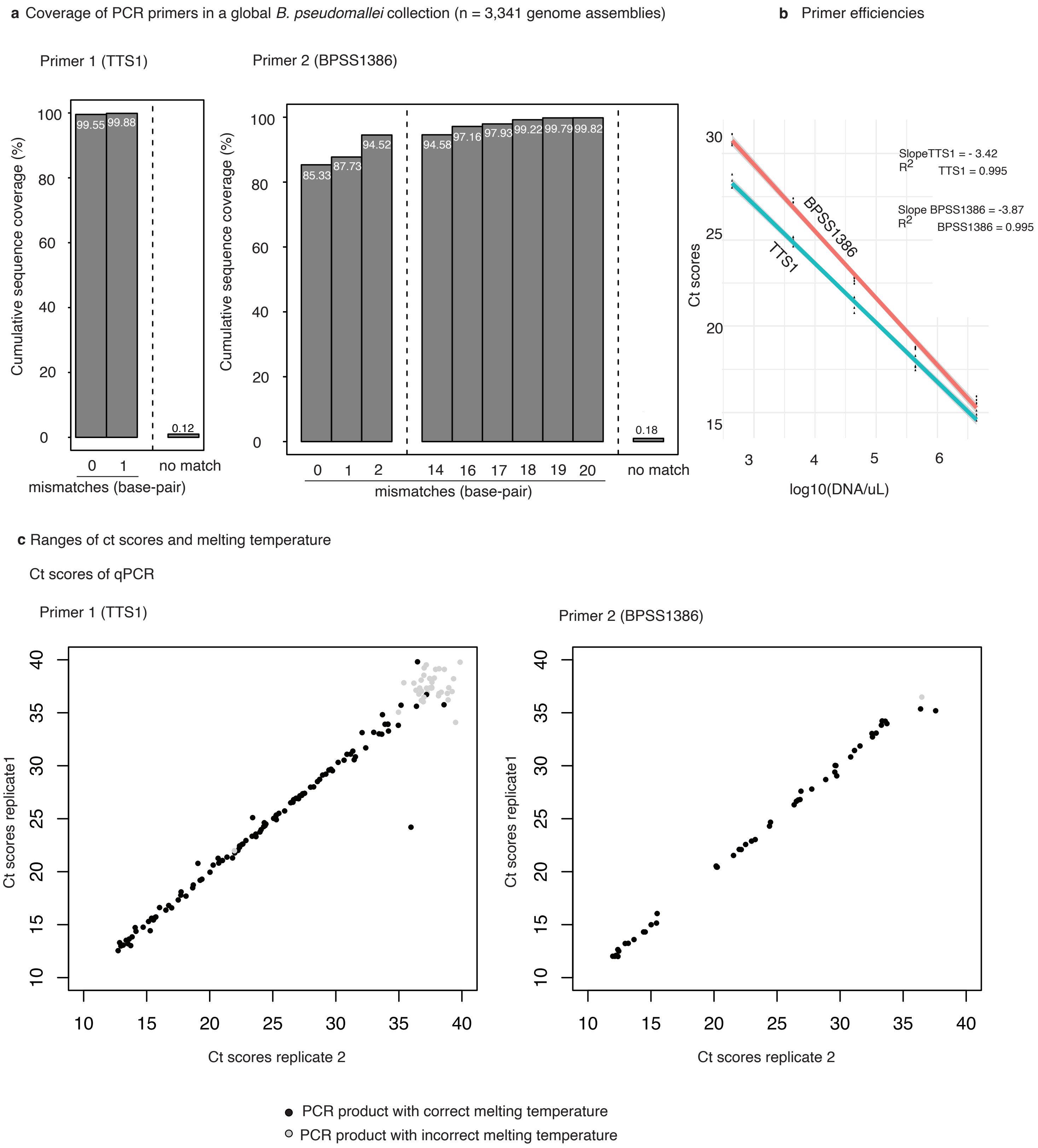

### Supplementary Tables

#### Table S1 A list of primers and oligos used in this study

| Name | Sequences | Usage | Reference |
|---|---|---|---|
| 148 | CAGCATATCATTGTCCGGCGCGAACCATCAAGCTA | RPA primer for crBP34 | [2] |
| 149 | AACTTTTCATTTTCCTGTCAATTCGACTGACCATC | RPA primer for crBP34 | [2] |
| 167 | TAATACGACTCACTATAGGGTAATTTCTACTAAGTG TAGATACTACATACCCACTATTCAGAAAGGAA | Annealing to DNA oligo 167 to create a T7transcription template for crBP34 crRNA | [2] |
| 168 | TTCCTTTCTGAATAGTGGGTATGTAGTATCTACACT TAGTAGAAATTACCCTATAGTGAGTCGTATTA | Annealing to DNA oligo 167 to create a T7transcription template for crBP34 crRNA | [2] |
| 192 | CGTCTCTATACTGTCGAGCAATCG | PCR primer targeting orf2 within putative type III secretion system cluster 1 (TTS1) of *B. pseudomallei* | [29] |
| 193 | CGTGCACACCGGTCAGTATC | PCR primer targeting orf2 within putative type III secretion system cluster 1 (TTS1) of *B. pseudomallei* | [29] |
| 270 | AACACTGACAAGTGGCCCTATGGA | PCR primer targeting orf11 (BPSS1386) of *B. pseudomallei* | [30] |
| 271 | TCCGATCGGTTTCGAATAACGGGT | PCR primer targeting orf11 (BPSS1386) of *B. pseudomallei* | [30] |
| FAM-biotin probe | /56-FAM/TTATT/3Bio/ | Collateral probe | [2,3] |

**Table S2 Demographic characteristics of the studied population**

| | Melioidosis cases (n=439 or stated otherwise) | Other infectious controls (n=190 or stated otherwise) | Healthy controls (n=506 or stated otherwise) |
|---|---|---|---|
| **Participant characteristics** | | | |
| Age, median years (IQR) | 54 (45-62) | 60 (52-69) | 50 (45-58) |
| Age groups, years (%) | | | |
| 18-30 | 22 (5%) | 7 (4%) | 0 |
| 31-50 | 154 (35%) | 36 (19%) | 275 (54%) |
| 51-70 | 225 (51%) | 110 (58%) | 214 (42%) |
| >70 | 37 (8%) | 36 (19%) | 17 (3%) |
| Sex (%) | | | |
| Female | 127 (29%) | 69 (37%) | 237 (47%) |
| Male | 311 (71%) | 120 (63%) | 269 (53%) |
| Self-reported ethnicity (%) | | | |
| Thai | 436 (>99%) | 187 (99%) | 504 (>99%) |
| Lao | 2 (<1%) | 2 (1%) | 1 (<1%) |
| Cambodian | 0 | 0 | 1 (<1%) |
| **Participant underlying health conditions** | | | |
| Body-mass index, median BMI scores (IQR) | 22 (20-25) | 22 (20-26) | 25 (23-28) |
| Body-mass index, BMI groups (%) | | | |
| Underweight (<18.5) | 52 (12%) | 23 (12%) | 13 (3%) |
| Healthy (18.5 – 22.9) | 192 (44%) | 78 (41%) | 109 (22%) |
| Overweight (23.0 – 26.5) | 139 (32%) | 51 (27%) | 220 (43%) |
| Obese (≥ 27.0) | 55 (12%) | 37 (19%) | 164 (32%) |
| unknown | 1 (<1%) | 1 (<1%) | 0 |
| Blood glucose level, HbA1c (IQR) | 8.65 (6.00-12.38) | 5.90 (5.10-7.68) | 6.45 (5.50-8.70) |
| Blood glucose level, groups | | | |
| Non-diabetic (<5.7) | 88 (20%) | 87 (46%) | 186 (37%) |
| Prediabetic (5.7 – 6.4) | 48 (11%) | 31 (16%) | 67 (13%) |
| Diabetic (≥ 6.5) | 298 (68%) | 72 (38%) | 253 (50%) |
| unknown | 5 (1%) | 0 | 0 |
| **Participant occupational and behavioural risks** | | | |
| Occupation (%) | | | |
| Agriculture and/or fisheries | 277 (63%) | 68 (36%) | 127 (25%) |
| Homemaker and/or retirees | 62 (14%) | 86 (46%) | 122 (24%) |

| | | | |
|---|---|---|---|
| Private sector | 55 (13%) | 16 (8%) | 128 (25%) |
| Merchant | 15 (3%) | 11 (6%) | 59 (12%) |
| Other | 29 (7%) | 8 (4%) | 69 (14%) |
| Exposure to flood in the past (%) | | | |
| True | 229 (52%) | 151 (80%) | 327 (65%) |
| False | 209 (48%) | 38 (20%) | 177 (35%) |

**Table S3 Univariate logistic regression of factors associated with melioidosis** (sample size = 1,132 participants)

| | **Odds ratio (95% CI)** | **p-value** |
|---|---|---|
| **Participants demographic** | | |
| Age | 1.00 (0.99-1.01) | 0.5 |
| Sex | | |
| Male (baseline) | 1 | |
| Female | 0.53 (0.41-0.70) | <0.001 |
| Self-reported ethnicity | | |
| Thai (baseline) | 1 | |
| Others | 0.70 (0.11-2.76) | 0.6 |
| **Participants underlying health conditions** | | |
| Body-mass index (BMI) | 0.88 (0.85-0.91) | <0.001 |
| Blood glucose level (HbA1c) | 1.22 (1.17-1.28) | <0.001 |
| **Participants occupational and exposure risks** | | |
| Occupation | | |
| Homemaker and/or retirees (baseline) | 1 | |
| Agriculture and/or fisheries | 5.48 (3.86-7.86) | <0.001 |
| Private sector | 1.28 (0.82-2.00) | 0.3 |
| Merchant | 0.71 (0.35-1.35) | 0.3 |
| Other | 1.27 (0.74-2.16) | 0.4 |
| Exposure to flood in the past 6 months | | |
| False (baseline) | 1 | |
| True | 2.02 (1.56-2.63) | <0.001 |

**Table S4 Multivariable logistic regression of factors associated with melioidosis based on *B. pseudomallei* detection within 10 km**
(sample size = 1,029 participants, 243 of which had direct sampling)

| | Risk of developing melioidosis | | | | | |
|---|---|---|---|---|---|---|
| | Environmental screening performed by conventional approach | | Environmental screening performed by CRISPR-BEEPs | | Environmental screening performed by double-primer qPCR | |
| | Odds ratio (95% CI) | p-value | Odds ratio (95% CI) | p-value | Odds ratio (95% CI) | p-value |
| **Participants demographic** | | | | | | |
| Age | 0.99 (0.98-1.00) | 0.2 | 0.99 (0.98-1.00) | 0.2 | 0.99 (0.98-1.00) | 0.2 |
| Sex | | | | | | |
| Male (baseline) | 1 | | 1 | | 1 | |
| Female | 0.57 (0.41-0.78) | <0.001 | 0.56 (0.41-0.78) | <0.001 | 0.56 (0.40-0.78) | <0.001 |
| Self-reported ethnicity | | | | | | |
| Thai (baseline) | 1 | | 1 | | 1 | |
| Others | 0.56 (0.07-2.69) | 0.5 | 0.55 (0.07-2.67) | 0.5 | 0.54 (0.06-2.57) | 0.5 |
| **Participants underlying health conditions** | | | | | | |
| Body-mass index (BMI) | 0.88 (0.84-0.91) | <0.001 | 0.88 (0.84-0.91) | <0.001 | 0.88 (0.84-0.91) | <0.001 |
| Blood glucose level (HbA1c) | 1.25 (1.19-1.32) | <0.001 | 1.25 (1.19-1.31) | <0.001 | 1.25 (1.19-1.31) | <0.001 |
| **Participants occupational and exposure risks** | | | | | | |
| Occupation | | | | | | |
| Homemaker and/or retirees (baseline) | 1 | | 1 | | 1 | |
| Agriculture and/or fisheries | 4.66 (3.05-7.21) | <0.001 | 4.47 (2.92-6.93) | <0.001 | 4.46 (2.91-6.91) | <0.001 |
| Private sector | 1.22 (0.72-2.07) | 0.5 | 1.24 (0.73-2.11) | 0.4 | 1.25 (0.73-2.12) | 0.4 |
| Merchant | 0.68 (0.31-1.41) | 0.4 | 0.71 (0.32-1.47) | 0.4 | 0.70 (0.32-1.45) | 0.3 |
| Other | 1.18 (0.63-2.20) | 0.6 | 1.21 (0.64-2.27) | 0.6 | 1.23 (0.65-2.30) | 0.5 |
| Exposure to flood in the past 6 months | | | | | | |
| False (baseline) | 1 | | 1 | | 1 | |
| True | 1.10 (0.79-1.52) | 0.6 | 1.09 (0.78-1.52) | 0.6 | 1.09 (0.78-1.51) | 0.6 |
| *B. pseudomallei* positivity rates in water samples within 10 km from the household | 1.03 (0.28-3.79) | >0.9 | 2.56 (1.31-5.04) | 0.006 | 2.74 (1.38-5.48) | 0.004 |

**Table S5 Multivariable logistic regression of factors associated with melioidosis based on *B. pseudomallei* detection within 9 km**
(sample size = 1,014 participants, 243 of which had direct sampling)

| | **Environmental screening performed by conventional approach** | | **Environmental screening performed by CRISPR-BEEPs** | | **Environmental screening performed by double-primer qPCR** | |
|---|---|---|---|---|---|---|
| | Odds ratio (95% CI) | p-value | Odds ratio (95% CI) | p-value | Odds ratio (95% CI) | p-value |
| **Participants demographic** | | | | | | |
| Age | 0.99 (0.98-1.00) | 0.2 | 0.99 (0.98-1.01) | 0.2 | 0.99 (0.98-1.00) | 0.2 |
| Sex | | | | | | |
| Male (baseline) | 1 | | 1 | | 1 | |
| Female | 0.55 (0.40-0.77) | <0.001 | 0.55 (0.39-0.77) | <0.001 | 0.55 (0.39-0.76) | <0.001 |
| Self-reported ethnicity | | | | | | |
| Thai (baseline) | 1 | | 1 | | 1 | |
| Others | 0.56 (0.07-2.73) | 0.5 | 0.57 (0.07-2.73) | 0.5 | 0.56 (0.07-2.66) | 0.5 |
| **Participants underlying health conditions** | | | | | | |
| Body-mass index (BMI) | 0.88 (0.85-0.91) | <0.001 | 0.88 (0.85-0.91) | <0.001 | 0.88 (0.84-0.91) | <0.001 |
| Blood glucose level (HbA1c) | 1.26 (1.20-1.32) | <0.001 | 1.25 (1.19-1.32) | <0.001 | 1.26 (1.20-1.32) | <0.001 |
| **Participants occupational and exposure risks** | | | | | | |
| Occupation | | | | | | |
| Homemaker and/or retirees (baseline) | 1 | | 1 | | 1 | |
| Agriculture and/or fisheries | 4.92 (3.20-7.66) | <0.001 | 4.77 (3.10-7.44) | <0.001 | 4.73 (3.07-7.39) | <0.001 |
| Private sector | 1.30 (0.76-2.23) | 0.3 | 1.33 (0.77-2.27) | 0.3 | 1.33 (0.78-2.28) | 0.3 |
| Merchant | 0.67 (0.29-1.43) | 0.3 | 0.69 (0.30-1.48) | 0.4 | 0.69 (0.30-1.47) | 0.4 |
| Other | 1.21 (0.63-2.28) | 0.6 | 1.23 (0.64-2.24) | 0.5 | 1.25 (0.65-2.47) | 0.5 |
| Exposure to flood in the past 6 months | | | | | | |
| False (baseline) | 1 | | 1 | | 1 | |
| True | 1.11 (0.80-1.55) | 0.5 | 1.13 (0.81-1.58) | 0.5 | 1.13 (0.81-1.58) | 0.5 |
| *B. pseudomallei* positivity rates in water samples within 9 km from the household | 1.10 (0.32-3.81) | 0.9 | 1.87 (1.01-3.50) | 0.048 | 2.11 (1.11-4.05) | 0.024 |

**Table S6 Multivariable logistic regression of factors associated with melioidosis based on *B. pseudomallei* detection within 8 km**
(sample size = 994 participants, 243 of which had direct sampling)

| | **Risk of developing melioidosis** | | | | | |
|---|---|---|---|---|---|---|
| | **Environmental screening performed by conventional approach** | | **Environmental screening performed by CRISPR-BEEPs** | | **Environmental screening performed by double-primer qPCR** | |
| | Odds ratio (95% CI) | p-value | Odds ratio (95% CI) | p-value | Odds ratio (95% CI) | p-value |
| **Participants demographic** | | | | | | |
| Age | 0.99 (0.98-1.01) | 0.2 | 0.99 (0.98-1.01) | 0.2 | 0.99 (0.98-1.00) | 0.2 |
| Sex | | | | | | |
| Male (baseline) | 1 | | 1 | | 1 | |
| Female | 0.56 (0.40-0.78) | <0.001 | 0.55 (0.40-0.77) | <0.001 | 0.55 (0.39-0.77) | <0.001 |
| Self-reported ethnicity | | | | | | |
| Thai (baseline) | 1 | | 1 | | 1 | |
| Others | 0.54 (0.06-2.63) | 0.5 | 0.57 (0.07-2.74) | 0.5 | 0.56 (0.07-2.68) | 0.5 |
| **Participants underlying health conditions** | | | | | | |
| Body-mass index (BMI) | 0.88 (0.84-0.91) | <0.001 | 0.88 (0.84-0.91) | <0.001 | 0.88 (0.84-0.91) | <0.001 |
| Blood glucose level (HbA1c) | 1.26 (1.20-1.33) | <0.001 | 1.26 (1.20-1.32) | <0.001 | 1.26 (1.20-1.32) | <0.001 |
| **Participants occupational and exposure risks** | | | | | | |
| Occupation | | | | | | |
| Homemaker and/or retirees (baseline) | 1 | | 1 | | 1 | |
| Agriculture and/or fisheries | 4.80 (3.12-7.49) | <0.001 | 4.70 (3.05-7.35) | <0.001 | 4.64 (3.01-7.26) | <0.001 |
| Private sector | 1.23 (0.72-2.12) | 0.4 | 1.25 (0.73-2.16) | 0.4 | 1.26 (0.73-2.16) | 0.4 |
| Merchant | 0.67 (0.29-1.43) | 0.3 | 0.68 (0.30-1.46) | 0.3 | 0.67 (0.29-1.44) | 0.3 |
| Other | 1.28 (0.66-2.42) | 0.5 | 1.29 (0.67-2.45) | 0.4 | 1.30 (0.68-2.46) | 0.4 |
| Exposure to flood in the past 6 months | | | | | | |
| False (baseline) | 1 | | 1 | | 1 | |
| True | 1.12 (0.81-1.59) | 0.5 | 1.17 (0.83-1.63) | 0.4 | 1.17 (0.83-1.63) | 0.4 |
| *B. pseudomallei* positivity rates in water samples within 8 km from the household | 2.33 (0.80-6.83) | 0.12 | 1.67 (0.93-3.00) | 0.89 | 1.96 (1.07-3.61) | 0.03 |

**Table S7 Multivariable logistic regression of factors associated with melioidosis based on *B. pseudomallei* detection within 7 km**
(sample size = 968 participants, 243 of which had direct sampling)

| | **Risk of developing melioidosis** | | | | | |
|---|---|---|---|---|---|---|
| | **Environmental screening performed by conventional approach** | | **Environmental screening performed by CRISPR-BEEPS** | | **Environmental screening performed by double-primer qPCR** | |
| | Odds ratio (95% CI) | p-value | Odds ratio (95% CI) | p-value | Odds ratio (95% CI) | p-value |
| **Participants demographic** | | | | | | |
| Age | 0.99 (0.98-1.01) | 0.4 | 0.99 (0.98-1.01) | 0.4 | 0.99 (0.98-1.01) | 0.4 |
| Sex | | | | | | |
| Male (baseline) | 1 | | 1 | | 1 | |
| Female | 0.55 (0.39-0.78) | <0.001 | 0.55 (0.39-0.77) | <0.001 | 0.55 (0.39-0.77) | <0.001 |
| Self-reported ethnicity | | | | | | |
| Thai (baseline) | 1 | | 1 | | 1 | |
| Others | 0.58 (0.07-2.79) | 0.5 | 0.59 (0.07-2.86) | 0.6 | 0.59 (0.07-2.81) | 0.5 |
| **Participants underlying health conditions** | | | | | | |
| Body-mass index (BMI) | 0.88 (0.84-0.91) | <0.001 | 0.88 (0.84-0.91) | <0.001 | 0.88 (0.84-0.91) | <0.001 |
| Blood glucose level (HbA1c) | 1.27 (1.20-1.33) | <0.001 | 1.26 (1.20-1.33) | <0.001 | 1.26 (1.20-1.33) | <0.001 |
| **Participants occupational and exposure risks** | | | | | | |
| Occupation | | | | | | |
| Homemaker and/or retirees (baseline) | 1 | | 1 | | 1 | |
| Agriculture and/or fisheries | 5.12 (3.30-8.96) | <0.001 | 5.05 (3.35-7.96) | <0.001 | 5.01 (3.22-7.91) | <0.001 |
| Private sector | 1.29 (0.74-2.22) | 0.4 | 1.30 (0.75-2.25) | 0.4 | 1.30 (0.75-2.26) | 0.3 |
| Merchant | 0.70 (0.30-1.50) | 0.4 | 0.71 (0.31-1.52) | 0.4 | 0.71 (0.31-1.53) | 0.4 |
| Other | 1.33 (0.69-2.55) | 0.4 | 1.36 (0.70-2.61) | 0.4 | 1.39 (0.71-2.67) | 0.2 |
| Exposure to flood in the past 6 months | | | | | | |
| False (baseline) | 1 | | 1 | | 1 | |
| True | 1.06 (0.75-1.50) | 0.7 | 1.07 (0.76-1.51) | 0.7 | 1.07 (0.76-1.50) | 0.7 |
| *B. pseudomallei* positivity rates in water samples within 7 km from the household | 1.50 (0.52-4.42) | 0.5 | 1.46 (0.85-2.52) | 0.02 | 1.91 (1.09-3.34) | 0.024 |

**Table S8 Multivariable logistic regression of factors associated with melioidosis based on *B. pseudomallei* detection within 6 km**
(sample size = 934 participants, 243 of which had direct sampling)

| | Risk of developing melioidosis | | | | | |
|---|---|---|---|---|---|---|
| | **Environmental screening performed by conventional approach** | | **Environmental screening performed by CRISPR-BEEPS** | | **Environmental screening performed by double-primer qPCR** | |
| | Odds ratio (95% CI) | p-value | Odds ratio (95% CI) | p-value | Odds ratio (95% CI) | p-value |
| **Participants demographic** | | | | | | |
| Age | 1.00 (0.98-1.01) | 0.6 | 1.00 (0.98-1.01) | 0.6 | 1.00 (0.98-1.01) | 0.6 |
| Sex | | | | | | |
| Male (baseline) | 1 | | 1 | | 1 | |
| Female | 0.57 (0.40-0.81) | 0.002 | 0.57 (0.40-0.80) | 0.002 | 0.56 (0.40-0.80) | <0.001 |
| Self-reported ethnicity | | | | | | |
| Thai (baseline) | 1 | | 1 | | 1 | |
| Others | 0.59 (0.07-2.91) | 0.6 | 0.61 (0.07-2.98) | 0.6 | 0.60 (0.07-2.92) | 0.6 |
| **Participants underlying health conditions** | | | | | | |
| Body-mass index (BMI) | 0.88 (0.85-0.92) | <0.001 | 0.88 (0.85-0.92) | <0.001 | 0.88 (0.85-0.92) | <0.001 |
| Blood glucose level (HbA1c) | 1.27 (1.20-1.33) | <0.001 | 1.26 (1.20-1.33) | <0.001 | 1.26 (1.20-1.33) | <0.001 |
| **Participants occupational and exposure risks** | | | | | | |
| Occupation | | | | | | |
| Homemaker and/or retirees (baseline) | 1 | | 1 | | 1 | |
| Agriculture and/or fisheries | 5.69 (3.61-9.11) | <0.001 | 5.64 (3.58-9.03) | <0.001 | 5.59 (3.55-8.96) | <0.001 |
| Private sector | 1.39 (0.79-2.45) | 0.3 | 1.40 (0.79-2.46) | 0.2 | 1.39 (0.79-2.45) | 0.2 |
| Merchant | 0.73 (0.31-1.63) | 0.5 | 0.74 (0.31-1.65) | 0.5 | 0.74 (0.31-1.64) | 0.5 |
| Other | 1.32 (0.66-2.61) | 0.4 | 1.34 (0.67-2.63) | 0.4 | 1.34 (0.67-2.64) | 0.4 |
| Exposure to flood in the past 6 months | | | | | | |
| False (baseline) | 1 | | 1 | | 1 | |
| True | 1.08 (0.76-1.54) | 0.7 | 1.09 (0.76-1.55) | 0.6 | 1.09 (0.76-1.54) | 0.6 |
| *B. pseudomallei* positivity rates in water samples within 6 km from the household | 1.25 (0.41-3.78) | 0.7 | 1.30 (0.78-2.16) | 0.30 | 1.45 (0.86-2.46) | 0.2 |

**Table S9 Multivariable logistic regression of factors associated with melioidosis based on *B. pseudomallei* detection within 5 km**
(sample size = 868 participants, 243 of which had direct sampling)

| | **Risk of developing melioidosis** | | | | | |
|---|---|---|---|---|---|---|
| | **Environmental screening performed by conventional approach** | | **Environmental screening performed by CRISPR-BEEPS** | | **Environmental screening performed by double-primer qPCR** | |
| | Odds ratio (95% CI) | p-value | Odds ratio (95% CI) | p-value | Odds ratio (95% CI) | p-value |
| **Participants demographic** | | | | | | |
| Age | 1.00 (0.98-1.01) | 0.6 | 1.00 (0.98-1.01) | 0.6 | 1.00 (0.98-1.01) | 0.6 |
| Sex | | | | | | |
| Male (baseline) | 1 | | 1 | | 1 | |
| Female | 0.58 (0.40-0.82) | 0.003 | 0.57 (0.39-0.82) | 0.003 | 0.57 (0.39-0.82) | 0.003 |
| Self-reported ethnicity | | | | | | |
| Thai (baseline) | 1 | | 1 | | 1 | |
| Others | 0.58 (0.07-2.84) | 0.5 | 0.59 (0.07-2.90) | 0.6 | 0.60 (0.07-2.91) | 0.6 |
| **Participants underlying health conditions** | | | | | | |
| Body-mass index (BMI) | 0.89 (0.85-0.92) | <0.001 | 0.89 (0.85-0.92) | <0.001 | 0.89 (0.85-0.92) | <0.001 |
| Blood glucose level (HbA1c) | 1.26 (1.19-1.33) | <0.001 | 1.26 (1.19-1.33) | <0.001 | 1.26 (1.19-1.33) | <0.001 |
| **Participants occupational and exposure risks** | | | | | | |
| Occupation | | | | | | |
| Homemaker and/or retirees (baseline) | 1 | | 1 | | 1 | |
| Agriculture and/or fisheries | 6.04 (3.77-9.82) | <0.001 | 5.99 (3.74-9.75) | <0.001 | 5.94 (3.71-9.67) | <0.001 |
| Private sector | 1.30 (0.73-2.32) | 0.4 | 1.31 (0.73-2.34) | 0.4 | 1.31 (0.73-2.34) | 0.4 |
| Merchant | 0.76 (0.32-1.69) | 0.5 | 0.78 (0.33-1.74) | 0.6 | 0.78 (0.33-1.75) | 0.6 |
| Other | 1.33 (0.65-2.68) | 0.4 | 1.36 (0.67-2.73) | 0.4 | 1.37 (0.67-2.76) | 0.4 |
| Exposure to flood in the past 6 months | | | | | | |
| False (baseline) | 1 | | 1 | | 1 | |
| True | 1.11 (0.76-1.60) | 0.6 | 1.12 (0.77-1.61) | 0.6 | 1.12 (0.76-1.65) | 0.6 |
| *B. pseudomallei* positivity rates in water samples within 5 km from the household | 1.24 (0.38-4.01) | 0.7 | 1.41 (0.83-2.40) | 0.2 | 1.64 (0.96-2.81) | 0.07 |

**Table S10 Multivariable logistic regression of factors associated with melioidosis based on *B. pseudomallei* detection within 4 km**
(sample size = 801 participants, 243 of which had direct sampling)

| | **Risk of developing melioidosis** | | | | | |
|---|---|---|---|---|---|---|
| | **Environmental screening performed by conventional approach** | | **Environmental screening performed by CRISPR-BEEPS** | | **Environmental screening performed by double-primer qPCR** | |
| | Odds ratio (95% CI) | p-value | Odds ratio (95% CI) | p-value | Odds ratio (95% CI) | p-value |
| **Participants demographic** | | | | | | |
| Age | 0.99 (0.97-1.01) | 0.2 | 0.99 (0.97-1.01) | 0.2 | 0.99 (0.97-1.01) | 0.2 |
| Sex | | | | | | |
| Male (baseline) | 1 | | 1 | | 1 | |
| Female | 0.60 (0.41-0.88) | 0.01 | 0.59 (0.40-0.87) | 0.009 | 0.66 (0.40-0.88) | 0.009 |
| Self-reported ethnicity | | | | | | |
| Thai (baseline) | 1 | | 1 | | 1 | |
| Others | 0.60 (0.07-2.95) | 0.6 | 0.60 (0.07-2.94) | 0.6 | 0.60 (0.07-2.93) | 0.6 |
| **Participants underlying health conditions** | | | | | | |
| Body-mass index (BMI) | 0.88 (0.84-0.92) | <0.001 | 0.88 (0.85-0.92) | <0.001 | 0.88 (0.84-0.92) | <0.001 |
| Blood glucose level (HbA1c) | 1.27 (1.20-1.34) | <0.001 | 1.27 (1.20-1.34) | <0.001 | 1.27 (1.20-1.34) | <0.001 |
| **Participants occupational and exposure risks** | | | | | | |
| Occupation | | | | | | |
| Homemaker and/or retirees (baseline) | 1 | | 1 | | 1 | |
| Agriculture and/or fisheries | 5.46 (3.35-9.08) | <0.001 | 5.41 (3.32-9.02) | <0.001 | 5.43 (3.32-9.02) | <0.001 |
| Private sector | 1.23 (0.67-2.26) | 0.5 | 1.25 (0.68-2.29) | 0.5 | 1.25 (0.68-2.28) | 0.5 |
| Merchant | 0.69 (0.28-1.59) | 0.4 | 0.70 (0.28-1.62) | 0.4 | 0.70 (0.28-1.60) | 0.4 |
| Other | 1.32 (0.65-2.72) | 0.4 | 1.35 (0.65-2.75) | 0.4 | 1.35 (0.65-2.75) | 0.4 |
| Exposure to flood in the past 6 months | | | | | | |
| False (baseline) | 1 | | 1 | | 1 | |
| True | 1.12 (0.76-1.65) | 0.6 | 1.12 (0.76-1.65) | 0.6 | 1.12 (0.76-1.65) | 0.6 |
| *B. pseudomallei* positivity rates in water samples within 4 km from the household | 0.98 (0.28-3.33) | >0.9 | 1.29 (0.75-2.21) | 0.4 | 1.24 (0.72-2.14) | 0.4 |

**Table S11 Multivariable logistic regression of factors associated with melioidosis based on *B. pseudomallei* detection within 3 km**
(sample size = 679 participants, 243 of which had direct sampling)

| | **Risk of developing melioidosis** | | | | | |
|---|---|---|---|---|---|---|
| | **Environmental screening performed by conventional approach** | | **Environmental screening performed by CRISPR-BEEPS** | | **Environmental screening performed by double-primer qPCR** | |
| | Odds ratio (95% CI) | p-value | Odds ratio (95% CI) | p-value | Odds ratio (95% CI) | p-value |
| **Participants demographic** | | | | | | |
| Age | 0.99 (0.97-1.01) | 0.2 | 0.99 (0.97-1.00) | 0.2 | 0.99 (0.97-1.01) | 0.2 |
| Sex | | | | | | |
| Male (baseline) | 1 | | 1 | | 1 | |
| Female | 0.75 (0.49-1.14) | 0.2 | 0.75 (0.49-1.14) | 0.2 | 0.75 (0.49-1.14) | 0.2 |
| Self-reported ethnicity | | | | | | |
| Thai (baseline) | 1 | | 1 | | 1 | |
| Others | 0.47 (0.02-2.58) | 0.5 | 0.45 (0.02-2.44) | 0.4 | 0.45 (0.02-2.48) | 0.4 |
| **Participants underlying health conditions** | | | | | | |
| Body-mass index (BMI) | 0.88 (0.84-0.92) | <0.001 | 0.88 (0.84-0.92) | <0.001 | 0.88 (0.84-0.92) | <0.001 |
| Blood glucose level (HbA1c) | 1.23 (1.16-1.31) | <0.001 | 1.24 (1.16-1.31) | <0.001 | 1.23 (1.16-1.31) | <0.001 |
| **Participants occupational and exposure risks** | | | | | | |
| Occupation | | | | | | |
| Homemaker and/or retirees (baseline) | 1 | | 1 | | 1 | |
| Agriculture and/or fisheries | 4.63 (2.71-8.06) | <0.001 | 4.58 (2.68-7.97) | <0.001 | 4.60 (2.69-7.99) | <0.001 |
| Private sector | 1.07 (0.56-2.04) | 0.8 | 1.07 (0.56-2.04) | 0.8 | 1.07 (0.56-2.03) | 0.8 |
| Merchant | 0.63 (0.23-1.55) | 0.3 | 0.64 (0.23-1.57) | 0.3 | 0.63 (0.23-1.56) | 0.3 |
| Other | 1.33 (0.62-2.80) | 0.5 | 1.35 (0.63-2.84) | 0.4 | 1.34 (0.63-2.82) | 0.4 |
| Exposure to flood in the past 6 months | | | | | | |
| False (baseline) | 1 | | 1 | | 1 | |
| True | 1.33 (0.86-2.05) | 0.2 | 1.32 (0.85-2.04) | 0.2 | 1.32 (0.85-2.04) | 0.2 |
| *B. pseudomallei* positivity rates in water samples within 3 km from the household | 0.87 (0.19-3.68) | 0.9 | 1.34 (0.74-2.44) | 0.3 | 1.20 (0.66-2.17) | 0.6 |

**Table S12 Multivariable logistic regression of factors associated with melioidosis based on *B. pseudomallei* detection within 2 km**

(sample size = 583 participants, 243 of which had direct sampling)

| | **Risk of developing melioidosis** | | | | | |
|---|---|---|---|---|---|---|
| | **Environmental screening performed by conventional approach** | | **Environmental screening performed by CRISPR-BEEPS** | | **Environmental screening performed by double-primer qPCR** | |
| | Odds ratio (95% CI) | p-value | Odds ratio (95% CI) | p-value | Odds ratio (95% CI) | p-value |
| **Participants demographic** | | | | | | |
| Age | 0.99 (0.97-1.01) | 0.2 | 0.99 (0.97-1.01) | 0.2 | 0.99 (0.97-1.01) | 0.2 |
| Sex | | | | | | |
| Male (baseline) | 1 | | 1 | | 1 | |
| Female | 0.64 (0.39-1.02) | 0.064 | 0.64 (0.39-1.02) | 0.063 | 0.64 (0.39-1.02) | 0.064 |
| Self-reported ethnicity | | | | | | |
| Thai (baseline) | 1 | | 1 | | 1 | |
| Others | 0.86 (0.03-9.54) | >0.9 | 0.86 (0.03-9.50) | >0.9 | 0.85 (0.03-9.27) | >0.9 |
| **Participants underlying health conditions** | | | | | | |
| Body-mass index (BMI) | 0.86 (0.81-0.90) | <0.001 | 0.86 (0.81-0.90) | <0.001 | 0.86 (0.81-0.90) | <0.001 |
| Blood glucose level (HbA1c) | 1.24 (1.16-1.33) | <0.001 | 1.24 (1.16-1.33) | <0.001 | 1.25 (1.16-1.35) | <0.001 |
| **Participants occupational and exposure risks** | | | | | | |
| Occupation | | | | | | |
| Homemaker and/or retirees (baseline) | 1 | | 1 | | 1 | |
| Agriculture and/or fisheries | 4.58 (2.49-8.58) | <0.001 | 4.57 (2.49-8.56) | <0.001 | 4.58 (2.49-8.57) | <0.001 |
| Private sector | 0.99 (0.48-2.00) | >0.9 | 0.99 (0.48-2.00) | >0.9 | 0.98 (0.48-1.99) | >0.9 |
| Merchant | 0.87 (0.30-2.23) | 0.8 | 0.87 (0.30-2.23) | 0.8 | 0.86 (0.30-2.23) | 0.8 |
| Other | 1.25 (0.54-2.84) | 0.6 | 1.26 (0.54-2.85) | 0.6 | 1.25 (0.54-2.82) | 0.6 |
| Exposure to flood in the past 6 months | | | | | | |
| False (baseline) | 1 | | 1 | | 1 | |
| True | 1.42 (0.87-2.32) | 0.2 | 1.42 (0.86-2.32) | 0.2 | 1.42 (0.87-2.33) | 0.2 |
| *B. pseudomallei* positivity rates in water samples within 2 km from the household | 0.95 (0.21-3.80) | >0.9 | 1.10 (0.58-2.08) | 0.8 | 0.96 (0.50-1.83) | >0.9 |

**Table S13 Multivariable logistic regression of factors associated with melioidosis based on *B. pseudomallei* detection within 1 km**
(sample size = 443 participants, 243 of which had direct sampling)

| | **Risk of developing melioidosis** | | | | | |
|---|---|---|---|---|---|---|
| | **Environmental screening performed by conventional approach** | | **Environmental screening performed by CRISPR-BEEPS** | | **Environmental screening performed by double-primer qPCR** | |
| | Odds ratio (95% CI) | p-value | Odds ratio (95% CI) | p-value | Odds ratio (95% CI) | p-value |
| **Participants demographic** | | | | | | |
| Age | 0.97 (0.95-1.00) | 0.04 | 0.99 (0.98-1.00) | 0.2 | 0.97 (0.95-1.00) | 0.04 |
| Sex | | | | | | |
| Male (baseline) | 1 | | 1 | | 1 | |
| Female | 0.57 (0.30-1.03) | 0.068 | 0.97 (0.95-1.00) | 0.046 | 0.57 (0.30-1.03) | 0.068 |
| Self-reported ethnicity | | | | | | |
| Thai (baseline) | 1 | | 1 | | 1 | |
| Others | 0.92 (0.04-9.82) | >0.9 | 1.00 (0.04-11.00) | >0.9 | 0.99 (0.04-10.8) | >0.9 |
| **Participants underlying health conditions** | | | | | | |
| Body-mass index (BMI) | 0.85 (0.80-0.90) | <0.001 | 0.85 (0.80-0.91) | <0.001 | 0.85 (0.80-0.91) | <0.001 |
| Blood glucose level (HbA1c) | 1.25 (1.15-1.35) | <0.001 | 1.25 (1.15-1.35) | <0.001 | 1.25 (1.15-1.35) | <0.001 |
| **Participants occupational and exposure risks** | | | | | | |
| Occupation | | | | | | |
| Homemaker and/or retirees (baseline) | 1 | | 1 | | 1 | |
| Agriculture and/or fisheries | 4.03 (1.91-8.72) | <0.001 | 4.04 (1.91-8.75) | <0.001 | 4.05 (1.91-8.79) | <0.001 |
| Private sector | 0.90 (0.38-2.08) | 0.8 | 0.94 (0.40-2.16) | 0.9 | 0.93 (0.40-2.15) | 0.9 |
| Merchant | 0.90 (0.25-2.79) | 0.9 | 0.89 (0.25-2.75) | 0.8 | 0.89 (0.25-2.73) | 0.8 |
| Other | 1.48 (0.57-3.75) | 0.4 | 1.25 (0.59-3.86) | 0.4 | 1.51 (0.59-3.83) | 0.4 |
| Exposure to flood in the past 6 months | | | | | | |
| False (baseline) | 1 | | 1 | | 1 | |
| True | 1.63 (0.90-2.93) | 0.11 | 1.66 (0.92-2.99) | 0.089 | 1.66 (0.92-2.99) | 0.09 |
| *B. pseudomallei* positivity rates in water samples within 1 km from the household | 1.81 (0.47-6.40) | 0.4 | 1.11 (0.57-2.15) | 0.8 | 1.00 (0.51-1.93) | >0.9 |
